# Human LUBAC deficiency leads to autoinflammation and immunodeficiency by dysregulation in TNF-mediated cell death

**DOI:** 10.1101/2022.11.09.22281431

**Authors:** Hirotsugu Oda, Kalpana Manthiram, Pallavi Pimpale Chavan, Shuichiro Nakabo, Hye Sun Kuehn, David B. Beck, Jae Jin Chae, Michele Nehrebecky, Amanda K. Ombrello, Tina Romeo, Natalie Deuitch, Brynja Matthíasardóttir, Jim Mullikin, Jennifer Stoddard, Julie Niemela, Holly Anderton, Kate E. Lawlor, Hiroyuki Yoshitomi, Dan Yang, Manfred Boehm, Jeremy Davis, Pamela Mudd, Davide Randazzo, Wanxia Li Tsai, Massimo Gadina, Mariana J. Kaplan, Junya Toguchida, Christian Mayer, Sergio D. Rosenzweig, Kazuhiro Iwai, John Silke, Bertrand Boisson, Jean-Laurent Casanova, Anand Rao, Najoua Lalaoui, Ivona Aksentijevich, Daniel L. Kastner

## Abstract

The linear ubiquitin assembly complex (LUBAC) consists of HOIP, HOIL1 and SHARPIN, and is essential for proper immune responses. Patients with HOIP and HOIL1 deficiencies present with severe immunodeficiency, autoinflammation and glycogen storage. In mice, the loss of *Sharpin* leads to severe dermatitis due to excessive cell death in keratinocytes. Here we report the first patient with SHARPIN deficiency, manifesting fever, arthritis, colitis, chronic otitis media and hepatic glycogenosis but unexpectedly, not associated with dermatologic manifestations. Mechanistically, fibroblasts and B cells from patients with all three LUBAC deficiencies showed attenuated canonical NF-κB response and propensity to apoptosis mediated by TNF superfamily members. Furthermore, the SHARPIN deficient patient showed substantial reduction of adenoidal germinal center B cell development. Treatment of the SHARPIN deficient patient with anti-TNF therapies led to complete clinical and transcriptomic resolution of autoinflammation. These findings underscore the critical role of LUBAC as a gatekeeper for apoptosis-mediated immune dysregulation in humans.

## INTRODUCTION

Monogenic systemic autoinflammatory diseases (SAIDs) are a group of heterogenous disorders characterized by dysregulation in innate immunity leading to overproduction of pro-inflammatory cytokines and chemokines ^1, 2^. The described disease mechanisms of SAIDs include hyperactivation of inflammasomes, type I interferon and NF-κB pathways. Recent κ studies highlighted the role of ubiquitin and TNF-mediated cell death pathways in the pathogenesis of autoinflammation, both in humans and in mice ^3^.

The linear ubiquitin assembly complex (LUBAC) is a trimeric complex consisting of HOIL1-interacting protein (HOIP), heme-oxidized IRP2 ubiquitin ligase-1 (HOIL1), and SHANK-associated RH domain interactor (SHARPIN). LUBAC mediates the conjugation of linear ubiquitin chains, also known as the Met1 ubiquitin chains, to various target molecules of the canonical NF-κB pathway and other signaling complexes ^4–6^. HOIP is the main catalytic subunit of LUBAC with an E3 ligase activity for linear ubiquitylation whereas SHARPIN and HOIL1 function as scaffold proteins, although recent reports suggest that HOIL1 also has an E3 ligase activity for mono-ubiquitylation ^7, 8^. Both HOIL1 and SHARPIN are necessary for LUBAC stability, and the depletion of any of these proteins greatly reduces the expression and the activity of LUBAC. Patients with HOIP and HOIL1 deficiencies carry biallelic loss-of-function (LOF) mutations in the respective genes and present with early-onset potentially lethal immunodeficiency, autoinflammation and glycogen deposits in the heart, skeletal muscle and liver ^9–11^. HOIP and HOIL1 deficient patients’ fibroblasts and B cells have signaling defects in the activation of the canonical NF-κB pathway, which accounts for the immunodeficiency phenotype. The autoinflammation was attributed to hyper-responsiveness of the blood cells, notably monocytes, to inflammatory cytokine stimulation. In mice, the ablation of HOIP (*Rnf31*) and HOIL1 (*Rbck1*) leads to embryonic lethality, wherea*s Sharpin* deficient mice are viable and present with severe TNF-dependent chronic proliferative dermatitis and multi-organ inflammation ^12^. LUBAC mutant mice have highlighted an essential contribution of apoptotic and necroptotic cell death to the inflammatory phenotypes seen in these mice ^12–14 15–17 18 19^. Nonetheless, the role of cell death pathways in human LUBAC deficiency is unknown.

This study describes the first patient with SHARPIN deficiency, termed *sharpenia*, who in stark contrast to patients with HOIL1 and HOIP deficiencies, presented with distinct clinical inflammatory features and with subtle immunodeficiency. In addition, the human phenotype is not reminiscent of the severe dermatologic phenotype in *Sharpin* deficient mice. Mechanistically, we show *ex vivo* and *in vivo* evidence that increased propensity to cell death, especially apoptosis mediated by TNF superfamily members, plays a major role in the pathogenesis of all three human LUBAC deficiencies, and that the inflammatory disease can be treated with anti-TNF therapy. These data provide a vindication of molecular medicine by underscoring a cardinal role of TNF-induced cell death in the pathogenesis of human LUBAC deficiency.

## RESULTS

### A patient with a homozygous LOF variant in *SHARPIN*

We studied a patient with a significant history of recurrent fever, parotitis, joint inflammation and colitis. At age between 0-5, he started having episodes of recurrent fever with severe painful parotid gland swelling (Fig. 1a), lymphadenopathy and hepatosplenomegaly. At age between 5-10, he developed swelling of his ankle (Fig. 1b and 1c) and knee joints, which progressively worsened to polyarthritis affecting the cervical spine (Fig. 1c), sacroiliac, and multiple other joints. He had frequent loose stools and two episodes of hematochezia. He did not present with any skin rash, and his skin biopsy for dermal fibroblast culture did not reveal histological evidence of inflammation. Given his clinical history, he was diagnosed with polyarticular juvenile idiopathic arthritis or chronic recurrent multifocal osteomyelitis. His treatment included steroids and methotrexate, without much improvement.

**Figure 1.**
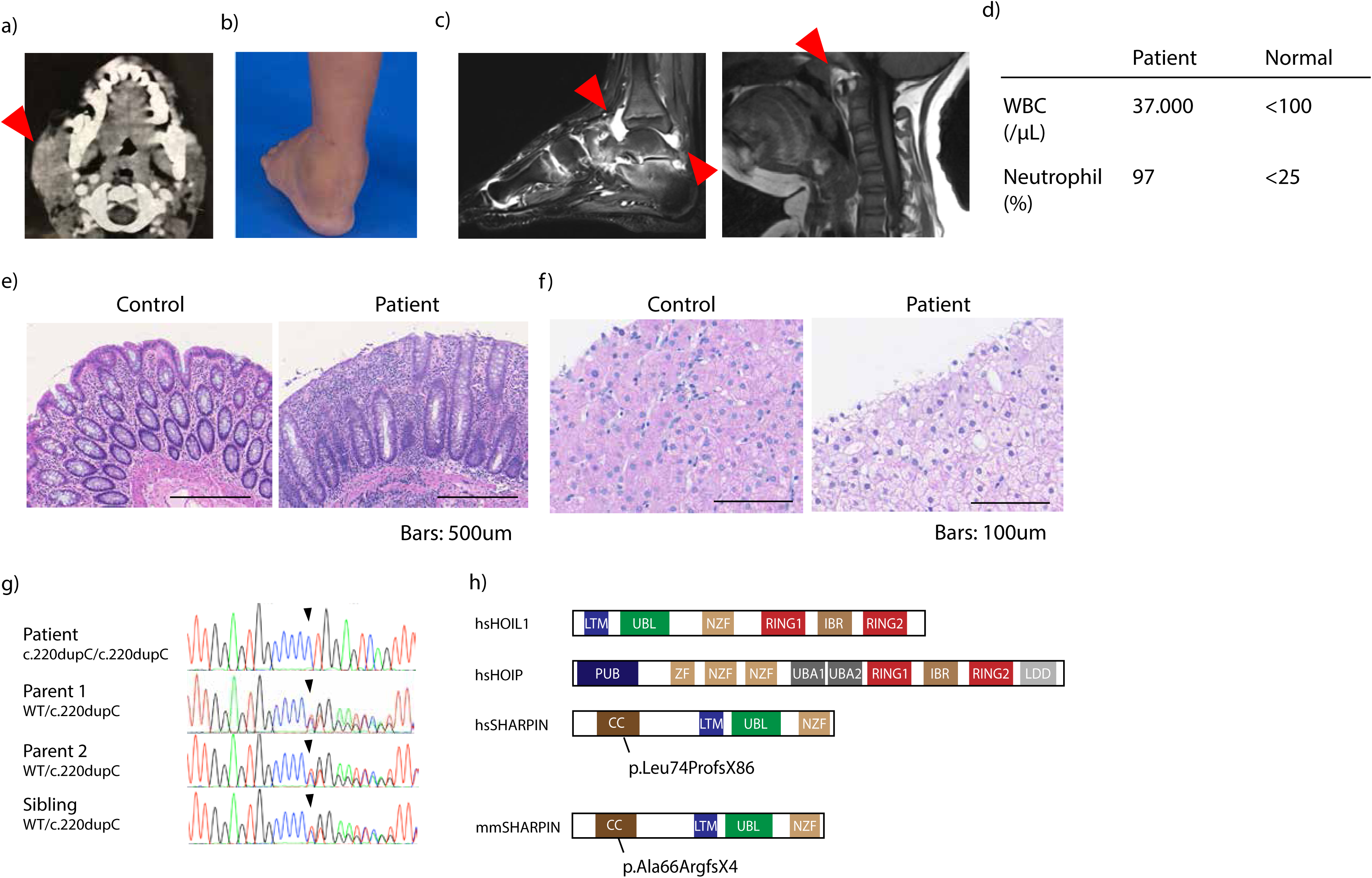
Human SHARPIN deficiency causes parotitis, polyarthritis, colitis and hepatic glycogenosis. (a) Computed tomography imaging demonstrating the swelling of the right parotid gland (arrowhead) at age between 0-5. (b) Swelling of the left ankle before initiation of treatment with etanercept. (c) Magnetic resonance imaging demonstrating joint inflammation of the ankle joint (left) and atlanto-axial joint (right). Arrowheads indicate inflammatory changes. (d) Pre-treatment sterile synovial fluid analysis from the ankle joint of SHARPIN deficient patient. (e) Colitis in SHARPIN deficient patient. (f) Hematoxylin and eosin staining of liver biopsy from SHARPIN deficient patient suggestive of glycogenosis. (g) Sanger sequence electropherograms demonstrating homozygous single base duplication in the patient. (h) Schematic domain structures of the three LUBAC subunits.

On initial examination at the National Institutes of Health at age between 10-15, he was wheelchair-dependent and had a prominent painful swelling of the left ankle joint (Fig. 1b). Magnetic resonance imaging scan detected high intensities in his ankle and atlanto-axial joints suggestive of active joint inflammation (Fig. 1c). Synovial fluid aspiration demonstrated marked leukocytosis with neutrophil predominance (Fig. 1d). Colon biopsy showed dense infiltration of lymphocytes and neutrophils in crypts and the lamina propria (Fig. 1e). Contrary to *Sharpin* deficient mice^20, 21^, there was no significant infiltration of eosinophils. Liver biopsy showed diffuse glycogen deposition (Fig. 1f), without inflammatory cells. Muscle biopsy was not performed due to the absence of myopathic clinical features. He experienced chronic group A streptococcal otitis media at age 15-20 with moderate conductive hearing loss, which necessitated tympanostomy tube drainage and adenoidectomy. No hypogammaglobulinemia was observed (IgG: 1506 mg/dL; IgA: 303 mg/dL; IgM: 243 mg/dL), and the IgG subclasses were normal (IgG1 717 mg/dL; IgG2: 508 mg/dL; IgG3: 79.3 mg/dL; IgG4: 49.4 mg/dL). Immunophenotyping of leukocyte surface markers did not show major abnormalities (Supplementary Data Table 1). Vaccine responses against PCV-13, VZV, HiB, diphtheria and tetanus were all positive (Supplementary Data Table 2). No autoantibodies were detected.

Given the patient’s early-onset severe inflammatory features and the parental consanguinity, we performed exome sequencing in the family. A stringent variant filtering for rare and novel homozygous variants (Extended Data Fig. 1a) identified a homozygous single base pair duplication in *SHARPIN* (NM_030974.4: c.220dupC) (Fig. 1g), which is predicted to induce a frameshift and a premature stop codon (NP_112236.3: p.Leu74Profs*86) (Fig. 1h). His asymptomatic parents and sibling were heterozygous carriers for the variant. This frameshift variant is not present in population genome databases (dbSNP, gnomAD and 1000 genomes) or in our in-house database of 1738 exomes from autoinflammatory patients. In silico analyses predicted this variant to be highly deleterious with a combined annotation-dependent deletion (CADD) score of 32.5, well above the mutation significance cutoff (MSC) score of 12.1 for SHARPIN (Extended Data Fig. 1b) ^22^. Furthermore, no homozygous LOF variants of *SHARPIN* or copy number loss encompassing *SHARPIN* in the homozygous state are reported in gnomAD ^23^, suggesting the intolerance to the complete loss of the gene.

### Impaired LUBAC expression and NF-κB activation in the SHARPIN deficient patient

The p.Leu74Profs*86 variant is in the NH2 terminal coiled-coil domain of the human SHARPIN, similar to the homozygous *Sharpin* variant (p.Ala66ArgfsX4) in *Sharpin* deficient mice that results in the complete loss of the SHARPIN protein (Fig. 1h) ^12–14, 24^. The patient’s PBMCs and fibroblasts did not show a detectable amount of the full-length SHARPIN protein, nor an expected 18kDa truncated protein (Fig. 2a-b). We also observed a significant reduction of HOIP expression, and in agreement with this, a reduced HOIP enrichment by HOIL1 immunoprecipitation was observed in the patient cells (Fig. 2c), which suggested the instability of the LUBAC. On the other hand, the mRNA expression of *SHARPIN* was reduced but not completely lost in the patient (Extended Data Fig. 2a-b). Furthermore, reconstitution of SHARPIN-KO HeLa cells with the patient-derived mutant protein did not produce a detectable amount of the truncated protein nor normalize the reduction of HOIP and HOIL1 expression (Extended Data Fig. 2c-d). These data indicate that the loss of SHARPIN protein in the patient cells is largely caused by the instability of the p.Leu74Profs*86 mutant SHARPIN protein.

**Figure 2.**
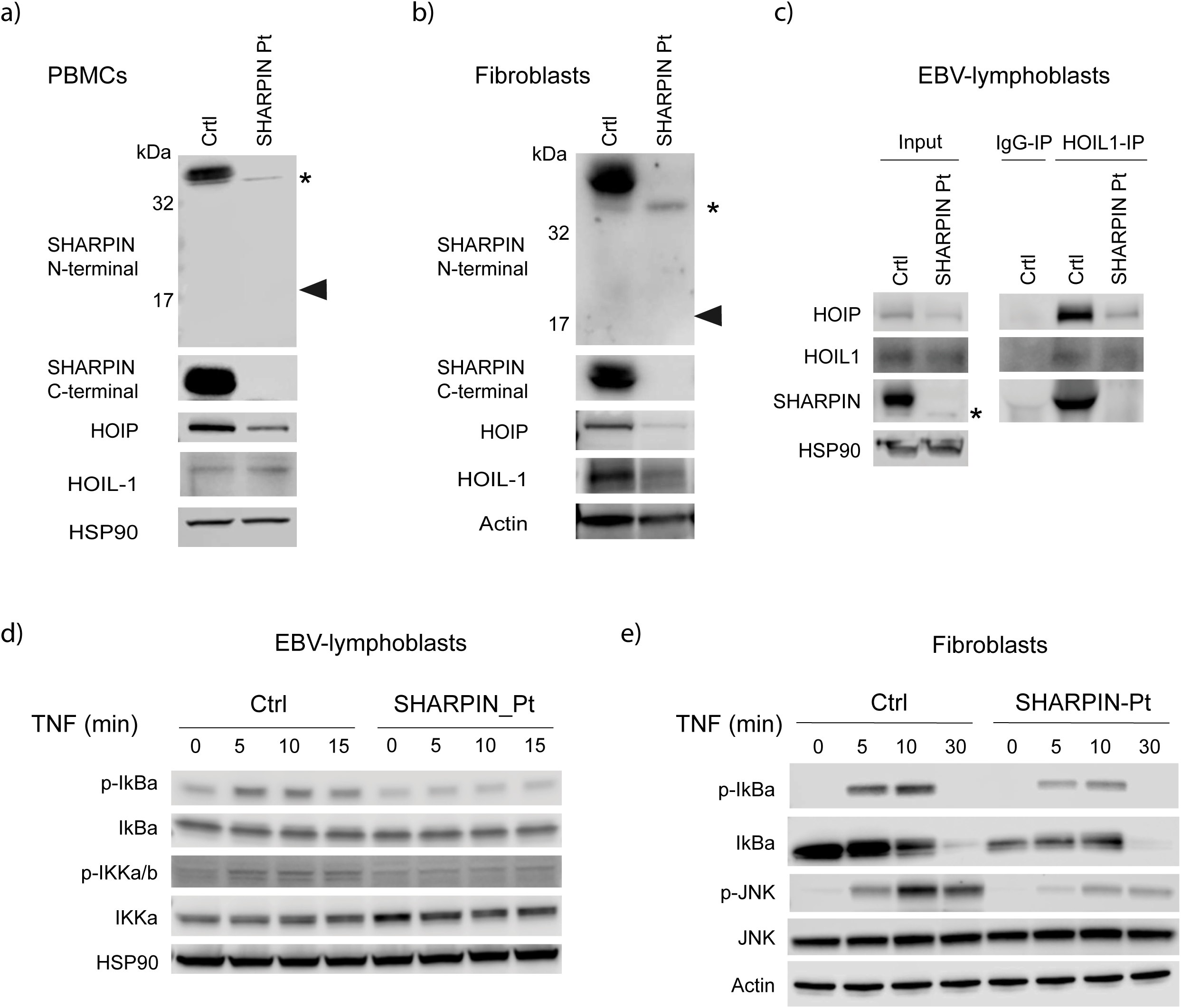
Human SHARPIN deficiency impairs canonical NF-B mediated signaling. (a-b) Protein expression of LUBAC subunits in (a) PBMCs and (b) fibroblasts. Note that the expected truncated SHARPIN protein (18kDa: arrowhead) was not observed. * indicates non- specific bands. (c) Western blot of immunoprecipitated extracts from EBV-immortalized lymphoblastic cells of healthy control and SHARPIN deficient patient. (d-e) SHARPIN deficient patient’s (d) EBV-immortalized lymphoblastic cells and (e) fibroblasts showed attenuated induction of NF-κB and MAPK after TNF stimulation (20 ng/ml), compared with an unrelated κ healthy control. \**p*<0.05, \*\*\**p*<0.001, *N.S*.: not significant (Student’s t-test). A representative result of three (a and d) or two (b, c and e) independent experiments is shown.

We next determined the effect of SHARPIN deficiency on TNF-induced NF- B κ activation in the patient fibroblasts. Consistent with previous reports in the *Sharpin* deficient mice ^13 12 14^, the SHARPIN deficient patient’s cells demonstrated reduced IkBα and JNK phosphorylation upon TNF stimulation, indicating the attenuation of NF- κB and MAPK activation (Fig. 2d and 2e). In contrast, CD3-induced non-canonical NF-κB activation was not affected in the patient cells (Extended Data Fig. 2e), consistent with a previous report in *Sharpin* deficient mice ^12, 13^. These data demonstrate that SHARPIN plays a role in canonical NF-κ B activation in humans.

### Increased induction of apoptosis in human LUBAC deficiencies

Previously, the inflammatory signature in human HOIL1 and HOIP deficiencies has been attributed to monocyte-derived inflammatory cytokines ^9, 10^. Here, we did not observe the higher cytokine production in stimulated monocytes from the SHARPIN deficient patient even during active disease (Extended Data Fig. 3). Failure to assemble plasma membrane-bound TNF receptor signaling complex (“complex I”) formation coincides with enhanced formation of a cytosolic RIPK1/FADD/caspase-8 complex (FADDosome) or RIPK1/RIPK3/MLKL complex (necrosome), which results in cell death by extrinsic apoptosis or necroptosis, respectively. Studies in LUBAC-deficient mice demonstrated that LUBAC is a gatekeeper of TNF-induced cell death by restricting the formation of complex II ^15–18^. We therefore tested whether the SHARPIN deficient patient cells were sensitized to TNF-induced cell death. In contrast to mouse *Sharpin* deficient cells ^17^, the patient fibroblasts did not undergo enhanced cell death when stimulated solely by TNF, whereas additional pharmacological manipulations such as smac mimetics (SM) or cycloheximide (CHX) increased the cell death induction *ex vivo* in the SHARPIN deficient patient cells compared to healthy control cells (Fig. 3a and b, and Extended Data Fig. 4a). We observed this propensity to cell death also in fibroblasts from patients with HOIP and HOIL1 deficiency as well as otulipenia and cleavage-resistant RIPK1-induced autoinflammation (CRIA), implicating that the cell death dysregulation may be a common feature underlying these human SAIDs.

**Figure 3.**
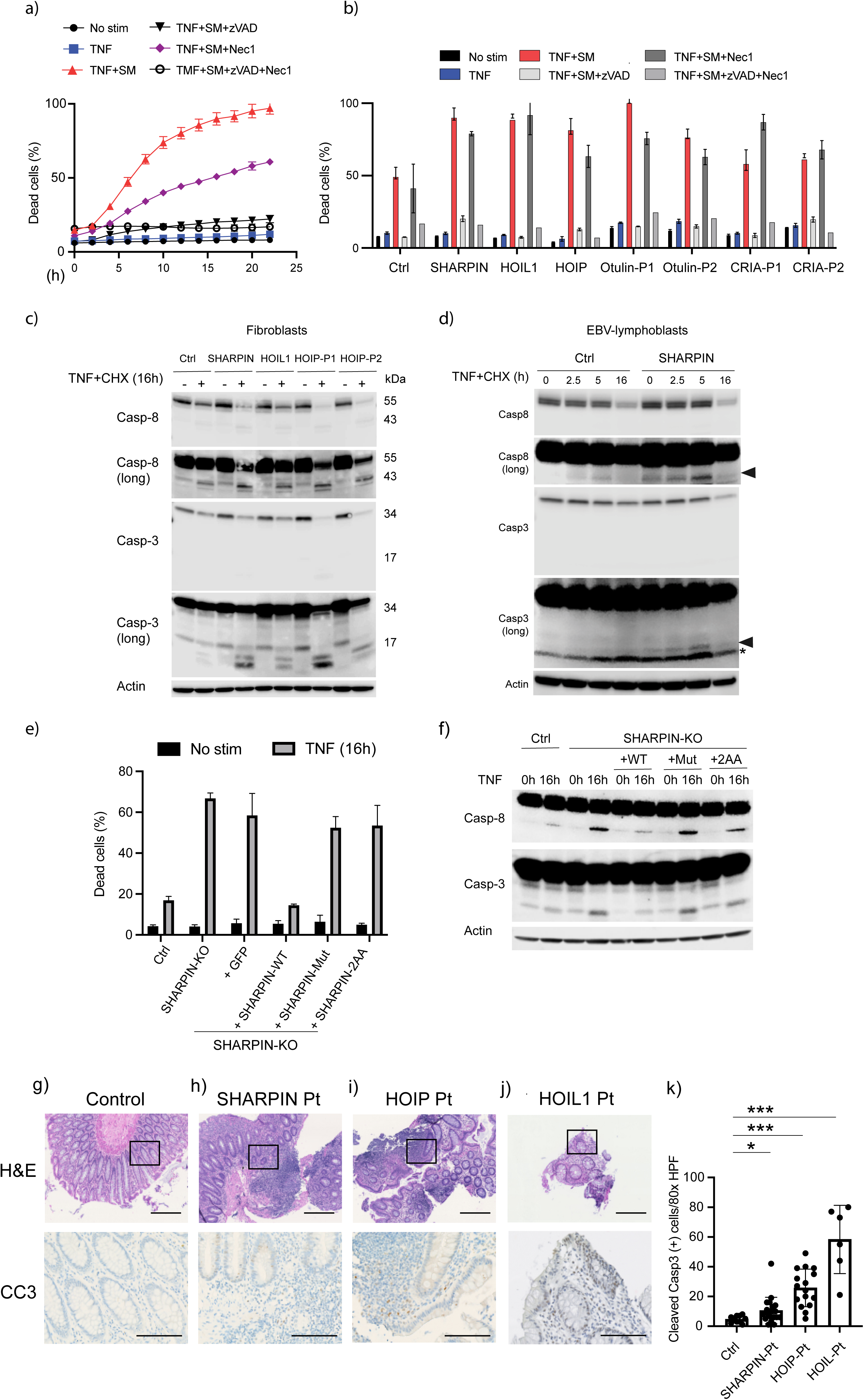
Human LUBAC deficiencies trigger excessive TNF-induced apoptosis. (a) Time-lapse quantification of dead cell percentages using fibroblasts from SHARPIN deficient patient. (b) Dead cell percentage at 20 h time point was quantified in fibroblasts from LUBAC deficiencies, otulipenia and cleavage resistant RIPK1-induced autoinflammation (CRIA). (a-b) The data acquisition was performed in biological triplicates. (c-d) Caspase cleavage assay in fibroblasts (c) and EBV-immortalized lymphoblasts (d) from LUBAC deficient patients. Long: long exposure. (e-f) Flow cytometry-based dead cell quantification (e) and caspase cleavage assay (f) using a SHARPIN-KO HeLa clone retrovirally reconstituted with GFP or SHARPIN variants. Mut: the frameshift variant observed in the patient (p.Leu74Profs*86); 2AA: a LUBAC-destabilizing positive control loss-of-function SHARPIN mutant (p.L179A/I183A) previously reported from structure-guided analysis ^19^. (a-f) The cells are stimulated with TNF (100ng/ml), cycloheximide (CHX:50 μ ml) or a smac mimetic (SM:compound A:100 nM) for the indicated time in the presence or absence of zVAD (pan-caspase inhibitor) or Nec1 (RIPK1 inhibitor). A representative result of three independent experiments is shown. (g-j) Histologic examination of colon biopsy samples from human LUBAC deficient patients. (k) Quantification of cleaved caspase-3 (CC3) positive cells per 80x high-power field (HPF). Bars indicate 400μm (upper panel) or 100μm (lower panel), respectively. \**p*<0.05, \*\*\**p*<0.001 (Student’s t-test).

Interestingly, in contrast to studies in mouse *Sharpin* deficient fibroblasts ^17^, this TNF-induced cell death was completely abolished by zVAD, a pan-caspase inhibitor, whereas it was partially suppressed by necrostatin-1, a RIPK1 inhibitor (Fig. 3a and b, and Extended Data Fig. 4a). Consistently with this cellular phenotype, we observed an augmented cleavage of apoptotic proteins caspase-3 and -8 in LUBAC deficient patients’ cells (Fig, 3c and 3d), whereas the phosphorylation of MLKL, a specific surrogate marker of necroptosis, was absent in EBV-immortalized B cells (EBV-B) and only minimally elevated in fibroblasts (Extended Data Fig. 4b and 4c). Thus, LUBAC deficient patients’ cells were largely sensitized to apoptosis with limited propensity to necroptosis, which contrasts with the previous reports from LUBAC-deficient mice as well as human cell death-mediated autoinflammatory disorders such as RIPK1 deficiency and TBK1 deficiency.

Next, to test the functional consequence of the p.Leu74Profs*86 frameshift SHARPIN variant, we examined the cell death in HeLa clones deficient in SHARPIN. Expectedly, SHARPIN deficient HeLa were susceptible to TNF-induced cell death primarily in a caspase-dependent manner, as confirmed by increased caspase-8 and -3 cleavage (Extended Data Fig. 4d and 4e). The TNF sensitivity in the SHARPIN deficient HeLa cells was not rescued by the overexpression of the mutant p.Leu74Profs*86 SHARPIN protein (Fig. 3e and 3f), further confirming the LOF nature of the frameshift variant.

Intestinal inflammation is a common disease feature in all three LUBAC- deficiencies. Indeed, colon biopsy samples from the LUBAC deficient individuals demonstrated a substantial infiltration of lymphocytes and neutrophils (Fig. 3g-j). Based on the increased cell death in the patient fibroblasts *ex vivo*, we investigated the extent of cell death *in vivo* by immunostaining for cleaved caspase-3 (CC3), which is a marker of apoptosis induction. The biopsy samples from the SHARPIN deficient patient demonstrated a moderate but significant increase in CC3 positivity mainly on intestinal crypts compared to a control sample (Fig. 3g and 3h). In contrast, colon samples from the HOIP- and HOIL1- deficient patients showed a more diffuse CC3 staining affecting crypts and interstitial areas (Fig. 3i-3k). Together, these *ex vivo, in vitro,* and *in vivo* data collectively demonstrate that heightened sensitivity to TNF-induced apoptosis contributes to the inflammatory phenotype in LUBAC deficient patients.

### Evidence of TNF-dependent inflammation in SHARPIN deficient joints

The SHARPIN deficient patient showed a significant sterile joint inflammation associated with massive neutrophil infiltration (Fig. 1b-d). To better characterize this disease feature, we measured the inflammatory cytokines in his synovial fluid. Compared to control samples with osteoarthritis, the SHARPIN deficient patient had a significantly higher level of IL6, whereas the level of TNF, known to accumulate in rheumatoid arthritis joints ^25^, was not elevated (Fig. 4a). The synovial fluid also showed significantly elevated levels of inflammatory chemokines (Fig. 4b and Extended Data Fig. 5a). Notably, we observed a prominent elevation of neutrophil chemoattractant proteins IL8 (CXCL8), GRO α (CXCL1), and MIP1α (CCL3) in agreement with the observed accumulation of neutrophils in the joint (Fig. 1d).

**Figure 4.**
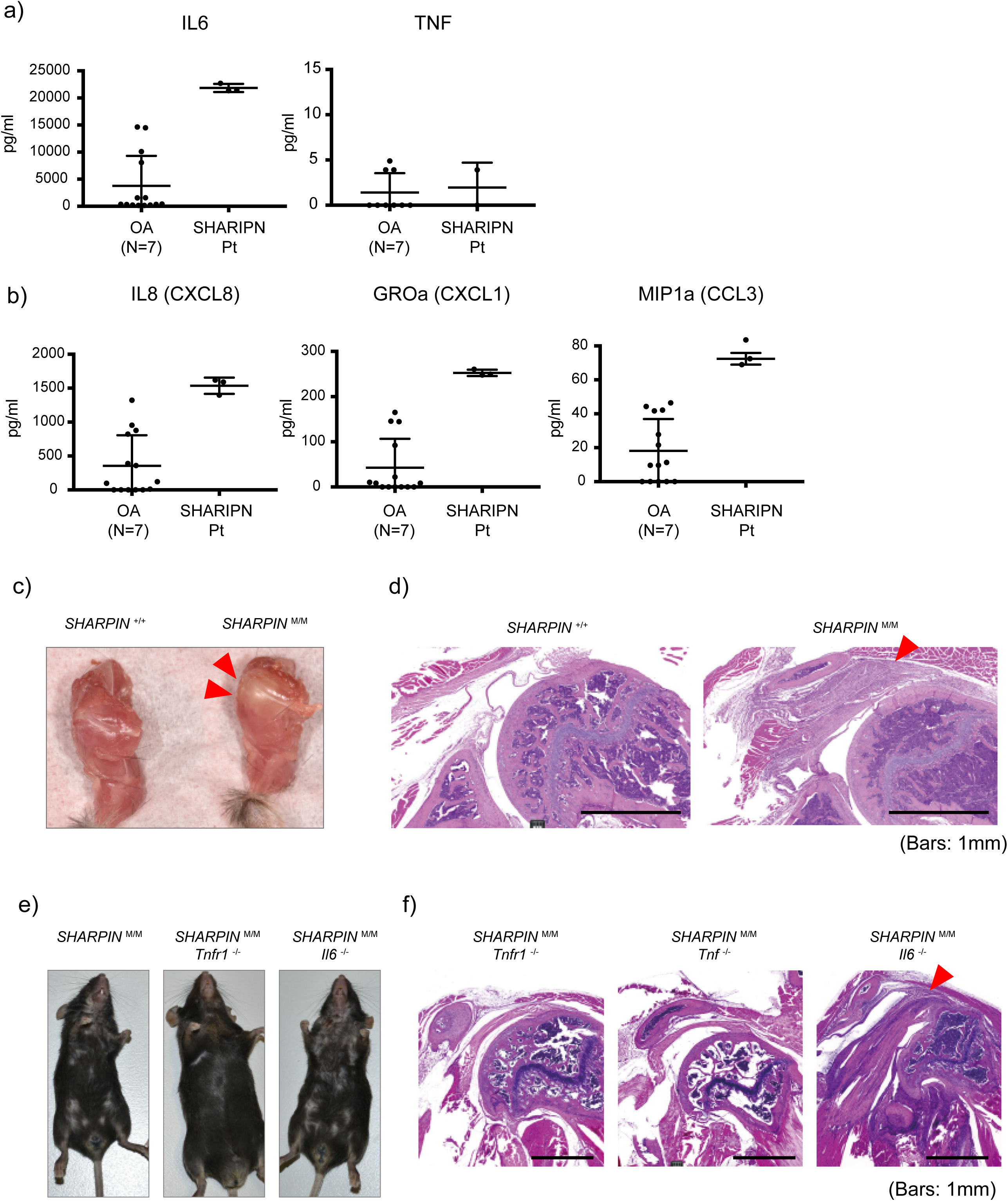
Loss of SHARPIN causes joint inflammation in human and mouse. (a-b) Multiplex ELISA measurement of (a) cytokines and (b) neutrophil-recruiting chemokines in the sterile synovial fluid of SHARPIN deficient patient before the initiation of anti-TNF treatment, compared with osteoarthritis (OA) controls (N=7). The samples were measured in technical triplicate (the SHARPIN deficient patient) or in duplicate (OA controls), respectively. (c) A representative photo of shoulder joints from *Sharpin* deficient mice (*Sharpin* ^M/M^) compared with wild type littermate control. Arrowheads indicate an inflamed shoulder joint capsule. (d) Representative hematoxylin and eosin (H&E) staining sections of shoulder joints from *Sharpin* deficient mice and wild type littermate control. Arrowhead indicates inflamed ligament indicative of enthesitis. (e) Representative dermatologic findings of *Sharpin* deficient mice crossed with *Tnfr1*- or *Il6*-KO mice. (f) Representative H&E staining sections of shoulder joints from *Sharpin* deficient mice crossed with *Tnf*-, *Tnfr1*- or *Il6*-KO mice.

Despite extensive studies of the *Sharpin* deficient mice which mostly focused on the skin inflammation, only two studies reported an arthritic phenotype in this animal model ^26^ ^27^; thus, the molecular histopathological mechanism of the joint inflammation in these mice remains unclear. Macroscopically, we observed a significant deposition of yellow plaque surrounding the shoulder joints of the *Sharpin* deficient mice (Fig. 4c), indicative of an inflammatory change. Histologically, this was accompanied by marked thickening of ligaments with massive immune cell infiltration, suggestive of enthesitis (Fig. 4d). This inflammatory finding was the most prominent on the shoulder, whereas a milder inflammatory finding was also observed on elbow joints (Extended Data Fig. 5b).

Of note, we did not observe histological evidence of chronic arthritis, such as erosions of bone surface or destruction of growth plates. Previous reports indicated an increased production of IL6 in monocytes from HOIL1 and HOIP deficient patients^9, 10^. Thus, we compared the contribution of IL6 signaling pathway with that of TNF on the inflammatory phenotype of *Sharpin* deficient mice by crossing them with *Il6*-, *Tnf*- and *Tnfr1*-KO mice. The deletion of *Tnf* or *Tnfr1* completely resolved the joint inflammation, concomitantly with their dermatitis as previously described ^12^ ^17^.

However, despite the accumulation of IL6 in HOIL1 and HOIP deficiet monocytes as well as the marked elevation of IL6 in the SHARPIN deficient patient’s synovial fluid, the deletion of *Il6* did not attenuate the inflammatory features of *Sharpin* deficient mice observed in their joints nor in the skin (Fig. 4e-f). These *in vivo* data collectively emphasize the importance of TNF-mediated inflammation on the pathology of joint inflammation in the SHARPIN deficient patient and its animal model.

### Impaired development of germinal centers in human LUBAC deficiencies

The SHARPIN deficient patient manifested relatively subtle features of immunodeficiency. This is in stark contrast to HOIP and HOIL1 deficiencies manifesting severe hypogammaglobulinemia, which has been in part explained by the attenuated B cell response to CD40L. To understand the molecular basis of this phenotypic difference, we dissected the histology of secondary lymphoid organs. In an axillary lymph node biopsy from one HOIP deficient patient ^11^, we observed remarkably few and disorganized lymphoid follicles (B cell zone). This finding was accompanied by no detectable germinal centers (GCs), highly specialized microanatomical sites with a role in B cell proliferation and maturation (Fig. 5a and Extended Data Fig. 6a).

**Figure 5.**
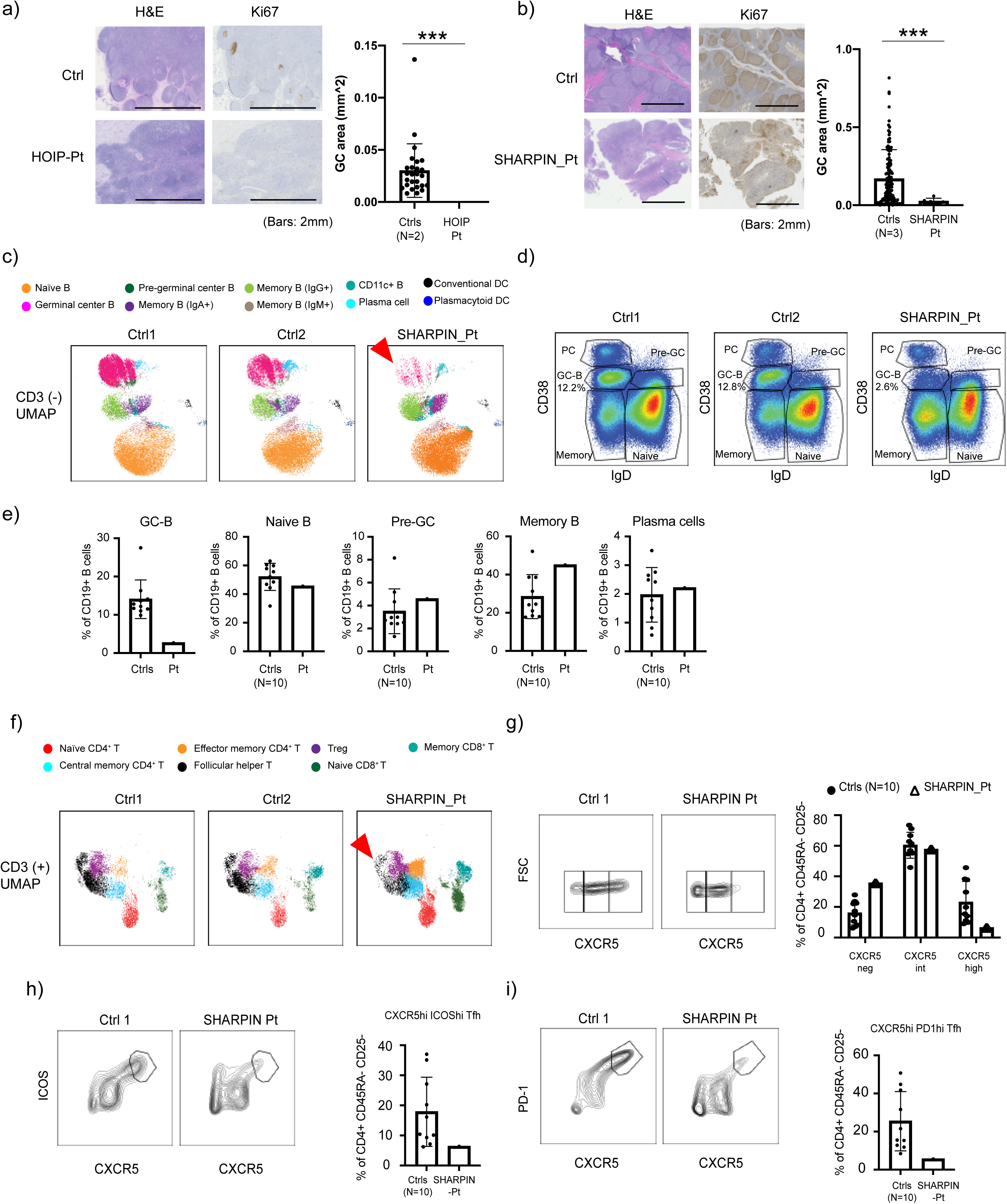
Human LUBAC deficiencies cause defective germinal center formation. (a) Lymph node histology of a HOIP deficient patient. (b) Adenoid histology of the SHARPIN deficient patient. (c) UMAP plot of CD3^-^population demonstrating reduction of germinal center B (GC-B) fraction (arrowhead) in SHARPIN deficient patient compared to controls (N=10). (d) Dot plot analysis of CD19^+^population with reduced CD19^+^CD38^int^IgD^-^GC-B in the patient. (e) Quantification of Fig. 6d. (f) UMAP plot of CD3^+^ population demonstrating reduction of follicular helper T cell fraction (arrowhead) in the SHARPIN deficient patient compared to controls (N=10). (g) Reduction of GC-Tfh (CD4^+^CD45RA^-^CD25^-^CXCR5^hi^) population in the SHARPIN deficient patient. (h-i) Reduced surface expression of ICOS (h) and PD1 (i) on GC-Tfh in the SHARPIN deficient patient. *** *p*< 0.001 (Student’s t-test).

Despite the milder immunodeficiency episodes, the SHARPIN deficient patient’s adenoids extracted during the treatment of otitis demonstrated very few, small and poorly structured lymphoid follicles and significantly reduced size of GCs (Fig. 5b and Extended Data Fig. 6b), consistent with a histological finding observed in B cell-specific *Sharpin* knock out mice ^28^. These data suggest that the LUBAC deficiency results in a dysregulated secondary lymphoid organ structure irrespective of the degree of clinical immunodeficiency.

Next, to investigate the effect of human SHARPIN deficiency on secondary lymphoid organ development, we established a novel 37-color high dimensional spectral flow cytometry assay using adenoid cells from the SHARPIN deficient patient and compared it with ten age-matched control samples. Consistent with the histology, we observed a reduction of the CD20^+^ B cell population in the patient (Extended Data Fig. 6c) along with a specific decrease in GC-B population (CD19^+^ CD38^int^IgD^-^) and a moderate increase of memory B cells (CD19^+^CD38^low^IgD^-^) (Fig. 5c-e). Despite the marked reduction of GC-B, relative class-switch recombination (CSR) was maintained in the GC-B and memory B cells (Extended Data Fig. 6d-e). This finding coincides with a previous report that CSR in B cells occurs before entering the GCs ^29^. Interestingly, the SHARPIN deficient patient’s adenoid had an intact percentage of CD19^+^ CD38^high^ IgD^-^plasma cell (PC) population (Fig. 5c-e). Recent studies have indicated that in secondary lymphoid organs, naïve B cells can differentiate into short-lived antibody-secreting cells through extrafollicular (EF) pathway independent of conventional GC incorporation, which contributes to rapid antibody response during infection ^30^. Based on the patient’s adenoidectomy during streptococcal otitis media, we reasoned that the normal percentage of adenoidal PC might be attributable to a potential activation of the EF pathway during the patient’s chronic infection.

We next focused on T cell populations in the adenoid analysis. Consistent with the histology (Extended Data Fig. 6b), we observed a moderate increase of CD3^+^T cells that can be attributed to increased proportions of central memory CD4^+^ and CD8^+^T cells (Extended Data Fig. 6f-g). Contrary to previous reports of murine *Sharpin* deficiency, ^31^ ^21^, we did not observe a reduction of the Treg population in the adenoid or in the peripheral blood (Extended Data Fig. 6f and Supplementary Data Table 1). T follicular helper cells (Tfh) are a specialized subset of CD4^+^T cells that support the appropriate formation of GCs and the subsequent production of high affinity immunoglobulin. Intriguingly, we observed an attenuated maturation of Tfh in the patient, demonstrated by the reduction in percentages of GC-follicular helper T cells (GC-Tfh: CD4^+^CD45RA^-^CD25^-^CXCR5^high^) (Fig. 5f and 5g). This was further accompanied by reduced expression of activation markers, ICOS and PD-1 (Fig. 5h-i). Altogether, these data indicate the importance of LUBAC in GC formation *in vivo*.

### Defective CD40L-induced B cell signaling in LUBAC deficiency *ex vivo*

To further gain insights into molecular mechanisms underlying the GC hypoplasia in the SHARPIN deficient patient *in vivo*, we examined the effect of the loss of SHARPIN on CD40-mediated B cell signaling, which reflects T cell-dependent B cell activation in GCs. The SHARPIN deficient patient’s B cells showed attenuated phosphorylation as well as target gene transcription of NF-κB and MAPK induced by CD40L (Fig. 6a-b). In agreement, CD40L-mediated transcription of activation-induced cytidine deaminase (AID), an activated B cell-restricted nucleotide-editing enzyme which underpins the class switching and affinity maturation of B cell receptors, was also attenuated in the SHARPIN deficient primary B cells (Fig. 6c). We further studied the consequence of the SHARPIN loss on cell fate. As shown in Fig. 6d and 6e, the SHARPIN deficient B cells had significant defects in cell survival associated with augmented cell death *ex vivo*. Importantly, we observed a more profound effect in a HOIP deficient patient compared with the SHARPIN deficient patient, implicating that these *ex-vivo* assays recapitulated the phenotypic difference between these two LUBAC deficiencies (Fig. 6c-e). We did not observe defects in T cell proliferation or Th1/2/17 skewing in contrast with previous reports of *Sharpin* deficient mice ^21^ ^32^, indicating that human SHARPIN may have a limited role in T cell homeostasis (Extended Data Fig. 7a-b). Lastly, we observed a higher degree of CC3-positive apoptotic cells among CD79^+^lymphoid follicles in SHARPIN- and HOIP-deficient secondary lymphoid organs *in vivo*, further highlighting the contribution of apoptosis dysregulations on the GC defects (Fig. 6f and 6g).

**Figure 6:**
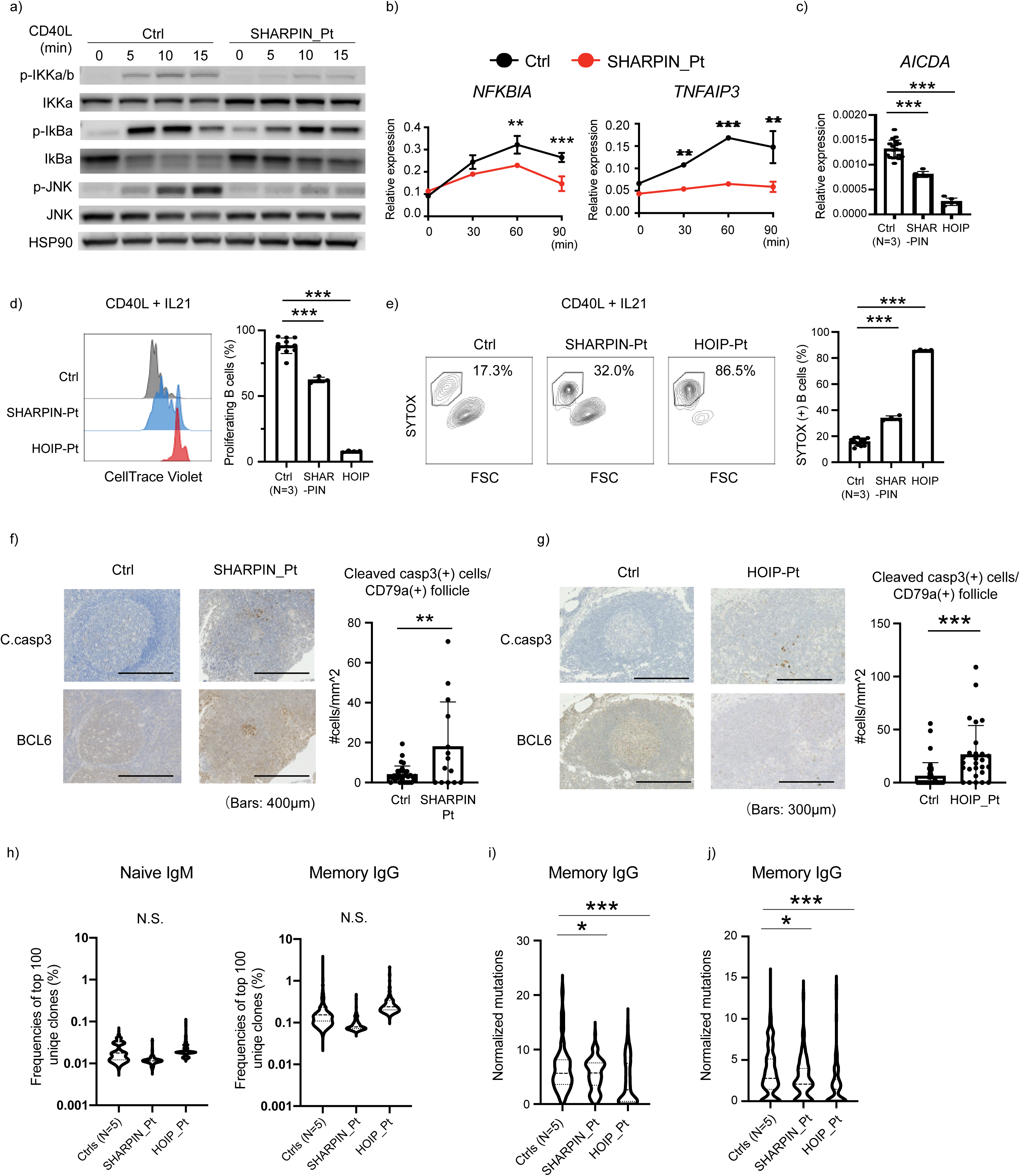
Human LUBAC deficiencies cause dysregulation in B cell activation and death. (a) NF-κB induction assay. EBV-immortalized lymphoblast (EBV-B) cells from SHARPIN κ deficient patient showed attenuated phosphorylation of IKKα/βI Bα and JNK after CD40L, stimulation. (b) mRNA expression of NF-κ stimulation. The experiment was performed in triplicates, and the expression levels were normalized to *GAPDH*. (c) mRNA expression of *AICDA* gene (encoding AID), normalized to *GAPDH*. CD19^+^primary B cells were enriched by anti-CD19 magnetic beads and stimulated with CD40L for 24h. The experiment was performed in quadruplicates. (d) Proliferation assay using primary B cells from LUBAC deficient patients. PBMCs were stained with CellTrace Violet, cultured with CD40L and IL21 for 96 h and analyzed by flow cytometer. (e) Cell death assay by SYTOX staining of primary B cells from LUBAC deficient patients. PBMCs were cultured with CD40L and IL-21 for 96 h. (f-g) Cleaved caspase-3 and BCL6 immunohistochemistry staining of (f) adenoid from SHARPIN deficient patient and (g) axillary lymph node from a HOIP deficient patient, compared with control tissues. The number of cleaved caspase-3 positive cells per follicle was normalized by CD79a^+^follicle area. (h-j) BCR sequencing from naïve and memory B cells, FACS-sorted from peripheral blood. (h) Relative frequencies of the 100 most abundant IGH clonotypes in LUBAC deficient patients and five age- matched healthy controls. (i-j) Somatic hypermutation (SHM) quantification in sorted B cells. SHM in the entire V region (i) and the CDR3 region (j) of the IGH gene were normalized by the nucleotide length of each clone. Normalized SHM of top 100 IGH clonotypes per each sample are demonstrated. * *p*<0.05, ** *p*<0.01, *** *p*< 0.001 (Student’s t-test). The data are representative of two (a-c) or three (d-e) independent experiments.

During the maturation process in the GCs, B cells undergo somatic hypermutation (SHM) to produce high-affinity clones. Indeed, patients with primary antibody deficiencies caused by dysfunction during GC maturation present with a wide range of abnormalities in the peripheral immunoglobulin repertoires ^33^_’_^34^. Thus, we investigated the role of LUBAC on immunoglobulin repertoires in the peripheral blood by targeted RNA sequencing of the BCR genes using sorted naïve and memory B cells from SHARPIN- and HOIP- deficient patients. There was no significant difference in the BCR clonality between the LUBAC deficient patients and the controls, indicating that the role of LUBAC on the global immunoglobulin repertoire is limited (Fig. 6h). In the memory B cells, however, we observed a significantly reduced SHM rate in the entire variable (V) region as well as the complementarity determining region 3 (CDR3) of IGHG gene (encoding IgG heavy chain), a hallmark of GC function to produce high affinity immunoglobulins *in vivo* (Fig. 6i and 6j). The SHM was clearly defective in the HOIP deficient patient, and was moderately attenuated in the SHARPIN deficient patient, indicating a genotype-phenotype correlation consistent with the *ex-vivo* functional data.

Collectively, our *ex vivo* and *in vivo* data indicate that the LUBAC-deficient patients had variable degrees of B cell immunodeficiency due to a dysfunction of germinal centers and propensity to cell death. Consequently, this defect led to the clinically severe immunodeficiency in the HOIP deficient patients, whereas the immunodeficiency phenotype of the SHARPIN deficient patient is less obvious.

### Genetics-guided treatment abolishes the inflammation in the SHARPIN deficient patient

Based on our functional data and the previously reported dependency of the systemic inflammation on TNF in the *Sharpin* deficient mice, we treated the SHARPIN deficient patient with etanercept, a recombinant soluble TNFR2-Fc fusion protein that antagonizes TNF. Notably, his arthritic symptoms remarkably improved within 1 month from the initiation of treatment, allowing him to ambulate without assistive devices (Fig. 7a). Because the patient still had intermittent GI inflammation, etanercept was switched to adalimumab, a monoclonal anti-TNF antibody indicated for inflammatory bowel disease. After the switch to adalimumab his colitis also markedly diminished (Fig. 7b), and the inflammatory markers were also completely normalized (Fig. 7c). This resolution of inflammatory disease was accompanied by a growth spurt (Fig. 7d) and full physical activity. There was also a substantial improvement of the patient’s bone mineral density (Fig. 7e), consistent with a report of TNF-dependent osteopenia in *Sharpin* deficient mice ^35^.

**Figure 7:**
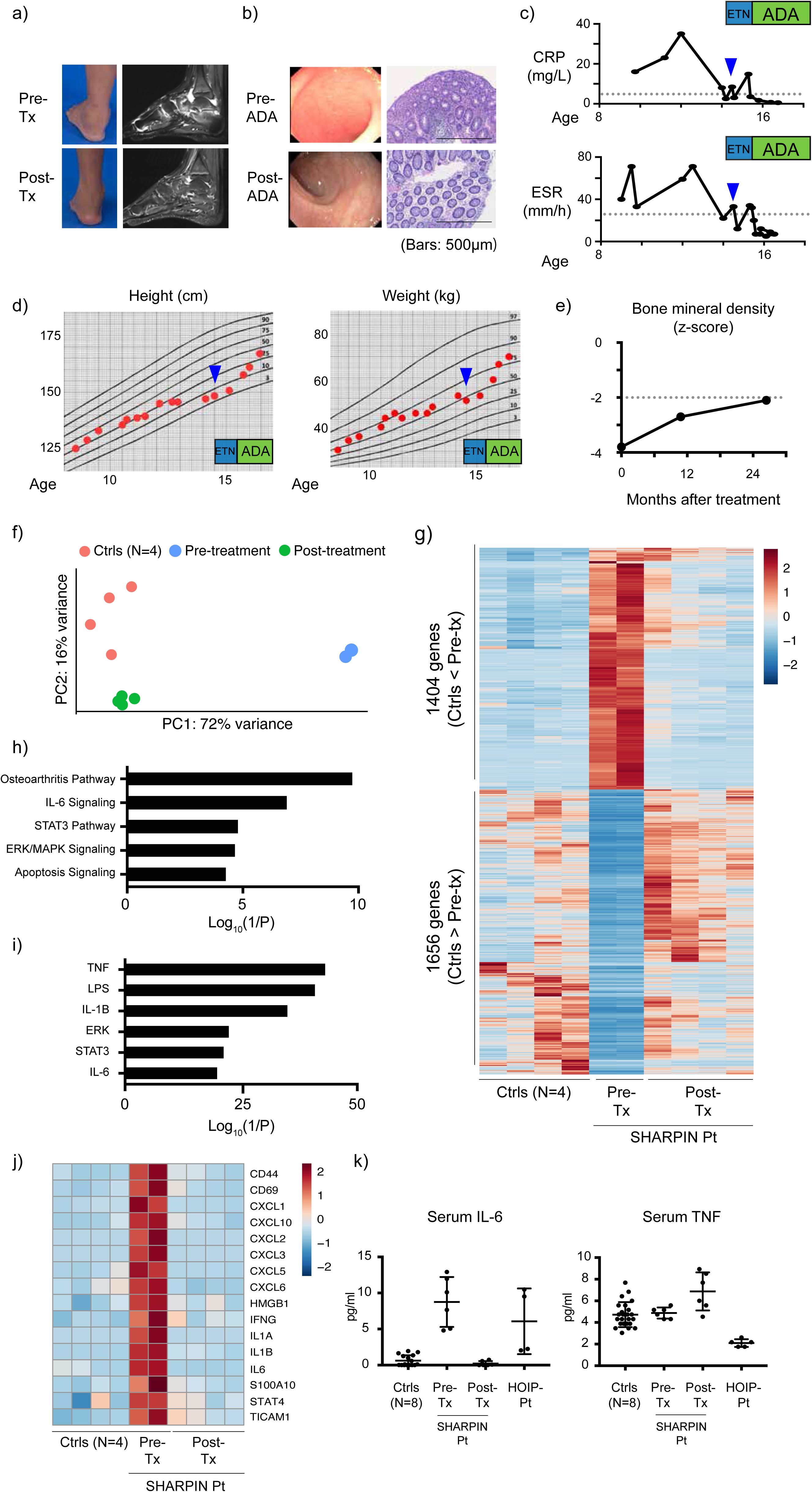
TNF biologics resolve the systemic inflammation of SHARPIN deficiency. (a-b) Clinical evaluation of (a) joint inflammation and (b) colonic inflammation to TNF blockade. (c) Response of inflammatory markers to TNF blocking therapies. (a-c) CRP: C-reactive protein; ESR: erythrocyte sedimentation rate; ETN: etanercept; ADA: adalimumab. (d) Growth recovery of SHARPIN deficient patient after TNF blockade. (e) Recovery of bone mineral density in SHRRPIN deficient patient after TNF blockade. (f) Principal component analysis using RNA sequencing data of pre- and post-treatment whole blood RNA samples from SHARPIN deficient patient. (g) A heatmap showing differentially expressed genes between healthy controls (N=4) and biological duplicate samples from pre-treatment SHARPIN deficient patient. (h-i) Gene enrichment analysis of 1404 genes upregulated in pre-treatment SHARPIN deficient patient using Ingenuity Pathway Analysis software. (h) Pathway analysis; (i) upstream molecule analysis. (j) A heatmap demonstrating the upregulation of representative inflammation-related genes in pre-treatment samples from SHARPIN deficient patient. (k) Response of serum IL-6 and TNF to TNF blockade therapies in SHARPIN deficient patient.

We further evaluated the effect of anti-TNF in the SHARPIN deficient patient at the transcriptomic level by whole blood RNA sequencing. The principal component analysis (PCA) clearly separated the pre-treatment data from healthy controls, whereas the post-treatment data approached the controls (Fig. 7f). The transcriptomic analysis identified 3060 differentially expressed genes (DEGs) between pre-treatment data and healthy controls (1404 DEGs upregulated and 1656 DEGs downregulated in the pre-treatment data). Consistent with the PCA, the expression of these DEGs were dramatically normalized in post-treatment data (Fig. 7g). By pathway analyses, we identified in the pre-treatment data an enrichment of multiple inflammatory pathways as well as upstream inflammatory molecules (Fig. 7h and 7i), including TNF-, IL1 β and IL6-mediated pathways. The increased inflammatory signature was markedly normalized by the anti-TNF treatment (Fig. 7j). Similarly, the cytokines and chemokines noted in the patient’s pre-treatment synovial fluid were significantly normalized in the post-treatment blood transcriptome, which include *IL6*, *CXCL8* (IL8), *CXCL1*(GROα), *CCL4* (MIP1β) and *CXCL10* (IP10) (Fig. 4a and 4b, Extended Data Fig. 8a and 8b). A serum ELISA assay validated the elevation of IL6 in the pre-treatment blood, which was normalized by anti-TNF (Fig. 7k). Interestingly, despite the patient’s complete response to anti-TNF, the protein and mRNA expression of TNF was not elevated in the pre-treatment blood (Fig.7k and Extended Data Fig. 8a). These clinical and functional data further support the hypothesis that the autoinflammation in human SHARPIN deficiency is caused by the augmented cell death of mutant cells due to their heightened responsiveness to TNF.

## DISCUSSION

In this study, we identified a novel recessively inherited human autoinflammatory disease, which we denote “*sharpenia*”. Remarkably, the patient presented with a phenotype distinct from patients with other LUBAC deficiencies and *Sharpin* deficient mice. Our *in vivo* and *ex vivo* analysis provide the first clinically relevant evidence for a role of human SHARPIN in autoinflammation, primarily driven by TNF-mediated apoptosis rather than necroptosis. Furthermore, despite the apparent lack of unusually severe infections in sharpenia we highlight a role of SHARPIN in human secondary lymphoid organ homeostasis by controlling B cell survival and death in GCs, induced by a second member of the TNF receptor superfamily, CD40. The remarkable efficacy of anti-TNF therapy in the sharpenia patient, supported by genetic dissection of joint inflammation using murine models, strongly suggests that TNF-mediated apoptosis contributes to the autoinflammation.

In the previous reports of human HOIL1 and HOIP deficiencies, the patients’ autoinflammation was attributed to a hyperresponsiveness of their monocytes to cytokine stimuli, leading to the increased production of IL6 and IL1β as well as inflammatory chemokines ^9, 10^. This upregulation was somewhat reminiscent of the accumulation of these inflammatory molecules in the blood and synovial fluid of sharpenia. In this study, however, we did not observe this cytokine hyperresponsiveness in sharpenia monocytes. Furthermore, our genetic dissection using *Il6* KO mice showed that the contribution of IL6 to autoinflammation is limited. This is further supported by recent publications indicating that the skin inflammation in *Sharpin* deficient mice is not fully resolved by the genetic ablation of IL1 signaling pathway, even though the expression of IL1β is highly elevated in their affected skin36 37 . In stark contrast, despite the unchanged TNF levels in sharpenia as well as in *Sharpin* deficient mice ^38^ ^39^, their autoinflammatory manifestations significantly improved upon TNF inhibition ^12^. Altogether, these findings highlight the critical importance of TNF-mediated signaling in driving autoinflammation in SHARPIN deficiency.

The inflammatory consequence of excessive cell death has been investigated in the past 20 years, primarily by elegant murine genetic studies. These animal models have provided a vast amount of evidence that excessive and unchecked programmed cell death, due to genetic, pharmacological or infectious insults, leads to uncontrolled inflammation *in vivo* ^40^ ^3^. The role of cell death-mediated inflammation has also been extensively studied in a number of human polygenic disorders, such as rheumatic and cardiovascular diseases, neuroinflammation and neurodegeneration, aiming to target the cell death pathways by inhibiting cell death effector kinases such as RIPK1 ^3^ ^41^.

However, those human patient-based studies are mainly restricted to correlative evidence, such as the upregulation of genes in cell death pathways (e.g., RIPK3) or histopathological characterizations. Recent advances in human genetics have enabled the rapid identification of causal genes in human SAIDs. For example, both LOF and gain-of-function mutations in RIPK1, a critical regulator of cell death signaling, have been reported in human SAIDs, indicating that the cell death-inducing activity of RIPK1 needs to be under strict regulatory control to ensure homeostasis in humans. Furthermore, other recently reported SAIDs, such as RELA haploinsufficiency, OTULIN deficiency and TBK1 deficiency, have provided additional causal genetic evidence of cell death-mediated inflammation in humans ^42–44^ ^45 46–48 49 50^. Here, we add LUBAC deficiency as a novel member of this clinical entity, which we propose to name “*inborn errors of cell death (IECD)*” (Supplementary Data Table 3).

The importance of studying IECD in humans, despite the recent availability of murine models, have been highlighted by the stark species-specific differences in phenotypes occasionally observed between human IECD and associated animal models. For example, in contrast to neonatal lethality in *Ripk1*-KO mice, patients with complete RIPK1 deficiency survive and manifest severe immunodeficiency with lymphopenia in addition to autoinflammation, implicating a critical role of RIPK1 on human adaptive immunity ^46^ ^45^. Here, the lack of dermatitis in sharpenia, without any trace of immune cell infiltration on histology, stands out as a novel dramatic example for this discrepancy.

Consistent with its severe skin inflammation, keratinocytes from *Sharpin* deficient mice were shown to be susceptible to TNF- and TLR3 agonist-induced cell death *ex vivo* ^16, 17^ ^51^, and their skin phenotype was rescued by crossing with either TNF haploinsufficient mice (*Tnf*^+/-^) or mice devoid of apoptotic and necroptotic machineries (*Casp8*^+/-^, *Casp8*^+/-^*Ripk3*^-/-^, *Casp8*^-/-^*Mlkl*^-/-^) ^12^ ^17 15^. Recently Sundberg et al., reported that conditional deletion of *Sharpin* in keratinocytes leads to severe dermatitis comparable to *Sharpin* deficient mice, whereas the skin phenotype of fibroblast-specific *Sharpin* KO mice is mild; in contrast, the severe arthritic phenotype is only observed in fibroblast-specific KO and not in keratinocyte-specific KO ^27^. Together with our patient data, these findings may indicate yet unknown cell lineage-specific mechanisms for the expression and function of LUBAC.

In this study, we observed that despite the normal lymphocyte markers in blood, the adenoidal lymphocytes from the sharpenia patient are developmentally affected. We observed a markedly defective GC formation in secondary lymphoid organs *in vivo*, caused by specific reduction of GC-associated lymphocytes, GC-B and GC-Tfh. We further provided mechanistic evidence by demonstrating the marked attenuation of CD40L-mediated activation and heightened sensitivity to cell death in LUBAC deficient patients’ B cells *ex vivo*, whereas the effect of human SHARPIN loss on T cell activation was seemingly limited. Recent reports have emphasized the indispensable role of GC-B in activating Tfh for their maturation and the subsequent collaborative formation of GC. Thus, we hypothesize that in sharpenia, the increased B cell death during GC maturation may be the initial cause of the GC hypoplasia. Lastly, we observed that the rate of SHM in LUBAC deficiencies *in vivo* correlates with the genotype and the degree of clinical immunodeficiency. Especially in sharpenia, the observed residual induction of AID, along with the intact PC potentially explained by compensatory EF differentiation, may be providing attenuated but still potent antibody production and affinity maturation. Recent human genetic studies have reported that patients with primary immunodeficiencies may manifest only after exposure later in life to specific pathogens including SARS-CoV2 ^52^, implicating a potential threshold for those immunodeficiencies to manifest clinically. A similar threshold may exist in antibody deficiencies, which could be detected in the form of avidity of antibodies ^53^. Whether the sharpenia patient as well as the HOIL1 deficient patients with non-lethal variants have functionally impaired antibodies needs to be further investigated in additional patients.

Beside their immunological manifestations, LUBAC deficient patients present with glycogen storage disease in skeletal muscle, heart and liver, although the molecular mechanism leading to this potentially life-threatening non-immune manifestation remains unclear. Recent work by Kelsall et al. demonstrated that mice with E3 ligase-inactive HOIL1 (p.Cys458Ser), devoid of immune dysregulation, develop spontaneous glycogenosis in the brain and the heart, reminiscent of LUBAC deficient patients ^54^.

They further provided mechanistic evidence that HOIL1, but not HOIP, directly ubiquitylates unbranched glucosaccharides *in vitro*, a novel paradigm-shifting example of ester-linked ubiquitylation of non-proteinaceous targets similarly to lipopolysaccharide^55^. These data are consistent with the clinical observation that HOIL1 deficient patients manifest with more severe glycogen storage phenotypes than HOIP deficient patients or the sharpenia patient. The glycogen accumulation in HOIL1 catalytic-inactive mice is age-dependent, especially in heart; thus, it is important to note that a careful follow-up is necessary for all LUBAC deficient patients who survive early-onset immunological complications.

In summary, we provided genetic and mechanistic evidence that human LUBAC deficiency results from the increased propensity to extrinsic apoptosis and consequently manifests with the two opposing forms of immune dysregulation, autoinflammation and B cell immunodeficiency. Studying the role of TNF-induced cell death in other human inborn errors of immunity will provide further insights for the development of targeted therapies.

## Data Availability

All data produced in the present study are available upon reasonable request to the authors

## ACKNOWLEDGEMENT

The authors would like to thank the patient and his family, and the healthy controls, for their enthusiastic support during this research study.

## METHODS

### Human subjects

All subjects gave written informed consent, including consent to publish, in accordance with the Declaration of Helsinki. This study is approved by the National Institutes of Health institutional review board (IRB), which serves as the central IRB for the study. **Exome sequencing (ES)** ES was performed using the Ion Torrent AmpliSeq RDY Exome kit (Life Technologies) and the Ion Chef and Proton instruments (Life Technologies). Briefly, 100 ng of gDNA was used as the starting material for the AmpliSeq RDY Exome amplification step following the manufacturer’s protocol. Library templates were clonally amplified and enriched using the Ion Chef and the Ion PI Hi-Q Chef kit (Chef package version IC.4.4.2; Life Technologies). Enriched, templated Ion Sphere Particles were sequenced on the Ion Proton sequencer using the Ion PI chip v3 (Life Technologies). Reads were mapped against the University of California, Santa Cruz (UCSC) hg19 reference genome using the Torrent Mapping Alignment Program (TMAP) map4 algorithm. Variants were called by the Torrent Variant Caller plugin (v.4.414-1) using the “Generic-Proton-Germ Line: Low Stringency” configuration. Variants were annotated using ANNOVAR (http://annovar.openbioinformatics.org/) ^56^.

### Whole blood RNA sequencing

Total RNA was isolated from whole blood collected in PAXgene Blood RNA tubes using PAXgene Blood RNA Kit (PreAnalytiX) per manufacturer’s instructions. Total RNA with high quality (RIN > 8) was used for cDNA library preparation using the NEBNext Ultra II Directional RNA library preparation kit with NEBNext Poly(A) mRNA magnetic isolation module and NEBNext Globin and rRNA Depletion kit (E7765, E7490 and E7755, New England Biolab). Sequencing was performed on an Illumina NovaSeq6000 System in a 2 x 150 bp paired-end mode. Sequenced reads were mapped against the human reference genome (GRCh38) using HISAT2 ^57^. Mapped reads were quantified using HTSeq ^58^. All the count data were normalized by total mapped reads, and differentially expressed genes were detected using edgeR ^59^. Pathway enrichment analysis was performed using Ingenuity Pathway Analysis (IPA) software (Qiagen).

### Plasmids and antibodies

The open reading frame of human SHARPIN gene (Addgene, #50014) was subcloned into retroviral vector (pDS-FB-IRES-hygromycin, kindly shared from Dr. Iain Fraser, NIAID, NIH) using Gateway system (Invitrogen). Mutant vectors are constructed using site-directed mutagenesis.

The following antibodies were used for immunoblotting: HOIP (Abcam: ab46322); HOIL1 (Santa Cruz:sc-393754); SHARPIN N-terminus (Millipore: ABF128); SHARPIN C-terminus (CST; 12541); β –Actin (Santa Cruz; sc-47778); IkBα (CST: 4814); p-IkB (CST: 2859); p-IKK α /β (CST: 2697); IKK α (CST: 11930); p-JNK (CST: 4668); JNK (CST: 9252); HSP90 (CST: 4877); Casp-8 (Enzo: ALX-804-242-C100); Casp-3 (CST: 9662); p-MLKL (CST:91689); MLKL (CST: 14993); p-RIPK1 (CST:65746); RIPK1 (CST: 3493).

### Cell culture, transfection and establishment of CRISPR-Cas9 knockout (KO) clones

Human embryonic kidney (HEK) 293T, HeLa and patient-derived SV40-immortalized fibroblasts ^9^ were maintained in DMEM with 10% fetal bovine serum and 1% PenStrep. Plasmid transfections were carried out using lipofectamine 2000 or NEON electroporation system (Invitrogen). LUBAC-deficient HeLa clones were established by transfecting CRISPR-Cas9 KO plasmid and HDR plasmid (Santa Cruz Biotech), followed by puromycin selection and single cell cloning. The status of target gene KO was confirmed by Western blot. Cells were routinely tested for mycoplasma using the Universal Mycoplasma Detection kit (ATCC).

### Western blotting and immunoprecipitation

Whole-cell lysates were prepared using ice-cold M-PER (Thermo Fisher Scientific) supplemented with protease inhibitor (Roche) and phosphatase inhibitor (Roche). Proteins were separated with Novex Tris-Glycine Gel Systems (Invitrogen) and transferred to nitrocellulose membranes with Trans-Blot Turbo systems (Biorad). After incubation with primary antibodies, proteins were visualized using ECL Plus Western blotting substrate (Thermo).

For LUBAC immunoprecipitation, cells were lysed using ice-cold lysis buffer containing 30 mM Tris-HCl, 150 mM NaCl, 1% Triton-X 100 and protease inhibitor cocktail (Roche). 1-2 mg of lysates were incubated with 3μg of anti-HOIL1 antibody (Santa Cruz: sc-393754) for 1 h, followed by incubation with protein A/G agarose beads for additional 30 min, washed 6 times with lysis buffer and subjected to immunoblotting.

### Time-lapse imaging-based cell death assay

SV40-immortaized fibroblasts were plated onto a 96-well plate (1×10^4/well, Corning #3596) and incubated overnight. The next day, the cells were stimulated and stained with SYTOX green (Thermo) and Nuclight Rapid Red Dye (Essenbioscience) for dead and live cells, respectively. Percentage of cell death was assayed every 1-2h by time-lapse imaging using the IncuCyte live cell analysis imaging (Essenbioscience) for 24h with 5% CO2 and 37 °C climate control.

### Histology and immunohistochemistry

Biopsies were fixed in formalin and embedded in paraffin before sectioning and staining.

Deparaffinization, hematoxylin and eosin (H&E) staining, and immunohistochemistry were performed by NDBbio Laboratories (Baltimore, MD). Stained slides were scanned, and image files were processed using NDP.view2 software (Hamamatsu photonics). Quantification was performed using ImageJ software ^60^.

### Flow cytometry

For intracellular cytokine studies, PBMCs were cultured with IL-1β (R/D, 10 ng/ml) and Monensin for 6 h and fixed with 4% PFA. Cells were permeabilized with Perm/Wash buffer (BD), immunolabeled with antibodies against CD14 (BD; 557154), TNF (BD; 554513), IL-6 (BD; 561441) and IL-1β (Abcam; 16168) and were analyzed by CytoFLEX (BD).

For proliferation assays, PBMCs were incubated with CellTrace Violet (Cell Proliferation Kit, Thermo) and stimulated with anti-IgM (Jackson Lab; 109-006-129), CD40L (Enzo; ALX522-110-C010), IL-21 (Peprotech; 200-21), CpG (Enzo; ALX522-025-C010), anti-CD3 (eBioscience; 16-0037-85), anti-CD28 (eBioscience; 16-0289-85) and PHA (Sigma; L9017). 72 or 96 h later the cells were stained with surface markers and analyzed by flow cytometer. Staining was performed with following antibodies from BD: CD3 (UCHT1), CD4 (RPAT4), CD8 (RPAT8), CD19 (HIB19) and CD80 (L307.4).

### Human adenoid analysis by high dimensional spectral flow cytometry

Adenoid cells were mechanically disrupted to make single cell suspension after adenoidectomy and cryopreserved until data acquisition. Cryopreserved adenoid cells were thawed and 5×10^6 cells per adenoid were used for the analysis. Cells were first stained with live dead dye for 15 min at room temperature (RT), washed twice and then incubated with true stain monocyte blocker (BioLegend) for 5 min. Antibodies for chemokine receptors (anti-CCR7 for 10 min, anti-CCR6, anti-CXCR5 and anti-CXCR3 together with brilliant stain buffer plus (BD) for 5min, anti-TCRγδ for 10 min) were sequentially added at RT. Antibody mix containing the rest of the surface antibodies and brilliant stain buffer plus were then added directly to the cells and incubated for 30 min at RT in the dark. Cells were washed three times and stained with fluorescence conjugated streptavidin for 15 min. Then cells were washed another three times and fixed in 1% paraformaldehyde for 20 min at RT before data acquisition on spectral flowcytometry (Aurora, Cytek). Data analysis was performed using Flowjo (version 10.7.2).

### Ex vivo cytokine secretion assay

Frozen PBMCs were resuspended in RPMI with 10% FBS and incubated for 1-2 h. After additional 30 min of serum starvation, the cells were further stimulated with LPS (1μg/ml) or IL-1β (10ng/ml) for 6 h. Cytokines in the supernatant were measured by ELISA (R&D Systems).

### Multiplexed detection of inflammatory cytokines and chemokines

Multiplex ELISA measurement of cytokines and chemokines in synovial fluid was performed using 34-Plex Human ProcartaPlex 1A (Thermo). The sample from the SHARPIN deficient patient was obtained before the initiation of anti-TNF treatment, and compared with osteoarthritis controls (N=7).

### 5’ RACE-based deep sequencing of the BCR genes

Total RNA was extracted from naïve and memory B cells (CD3^-^CD19^+^ CD27^-^and CD3^-^CD19^+^CD27^+^, respectively), sorted from bulk PBMCs of the SHARPIN and HOIP deficient patients as well as 5 healthy age-matched donors. 5’ RACE-based cDNA synthesis was performed according to the manufacture’s protocol (SMARTer Human BCR IgG/IgM H/K/L Profiling Kit, Takara Bio). Briefly, 10 ng of total RNA from sorted B cells were reverse transcribed using 5’RACE, with primers each contained a unique 8 bp unique molecular identifier (UMI) barcode sequences for demultiplexing. Synthesized cDNA was further amplified using multiplex PCR primers specific for IGHM and IGHG sequences. Sequencing was performed on an Illumina MiSeq system in a 2 x 300 bp paired-end mode. After demultiplexing UMI barcode sequences with MIGEC ^61^, the sequence reads were mapped using MiXCR ^62^. Immunoglobulin isotype-specific quantification of somatic hypermutation was conducted using MIGMAP ^61^. Downstream data analysis was performed using VDJtools ^63^.

### Animal experiment

The WEHI Animal Ethics Committee approved all mouse experiments, which were conducted according to the Australian Code for the care and use of animals for scientific purposes. All strains are on a C57BL/6 background with a minimum of five generation backcross.

### Statistical analysis

Statistical analyses were performed using GraphPad Prism (version 9.0.2)

## FIGURE LEGENDS

**Extended Data Figure 1:**
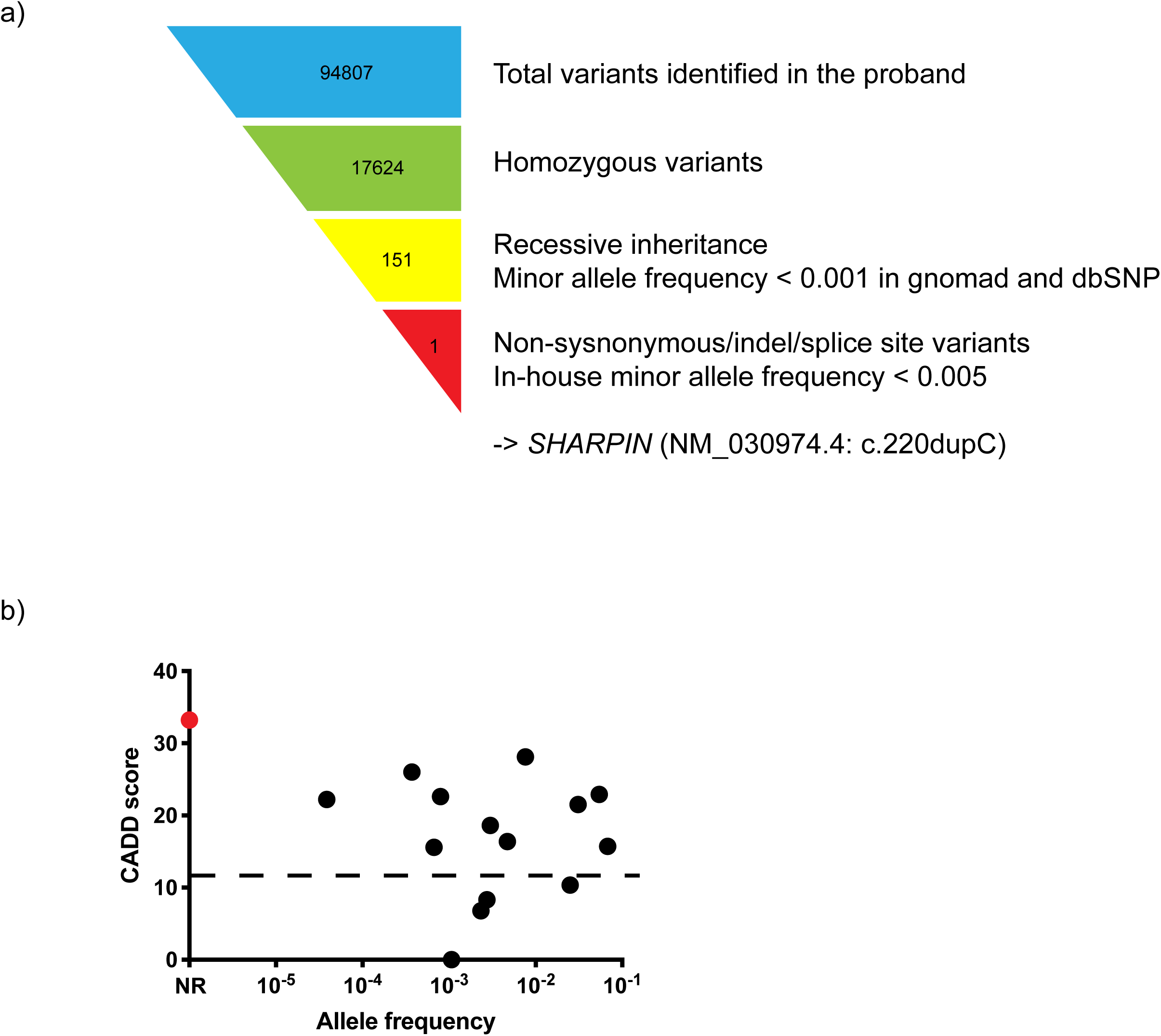
Genetic analysis of the pedigree. (a) Schematic representation of exome sequencing data processing. (b) Population allele frequency and CADD score for SHARPIN variants homozygous in public databases. The private SHARPIN variant c.220dupC appears in red. CADD-Mutation Significance Score (MSC) cutoff for SHARPIN (90% confidence interval) was indicated by dashed line. NR: not reported.

**Extended Data Figure 2:**
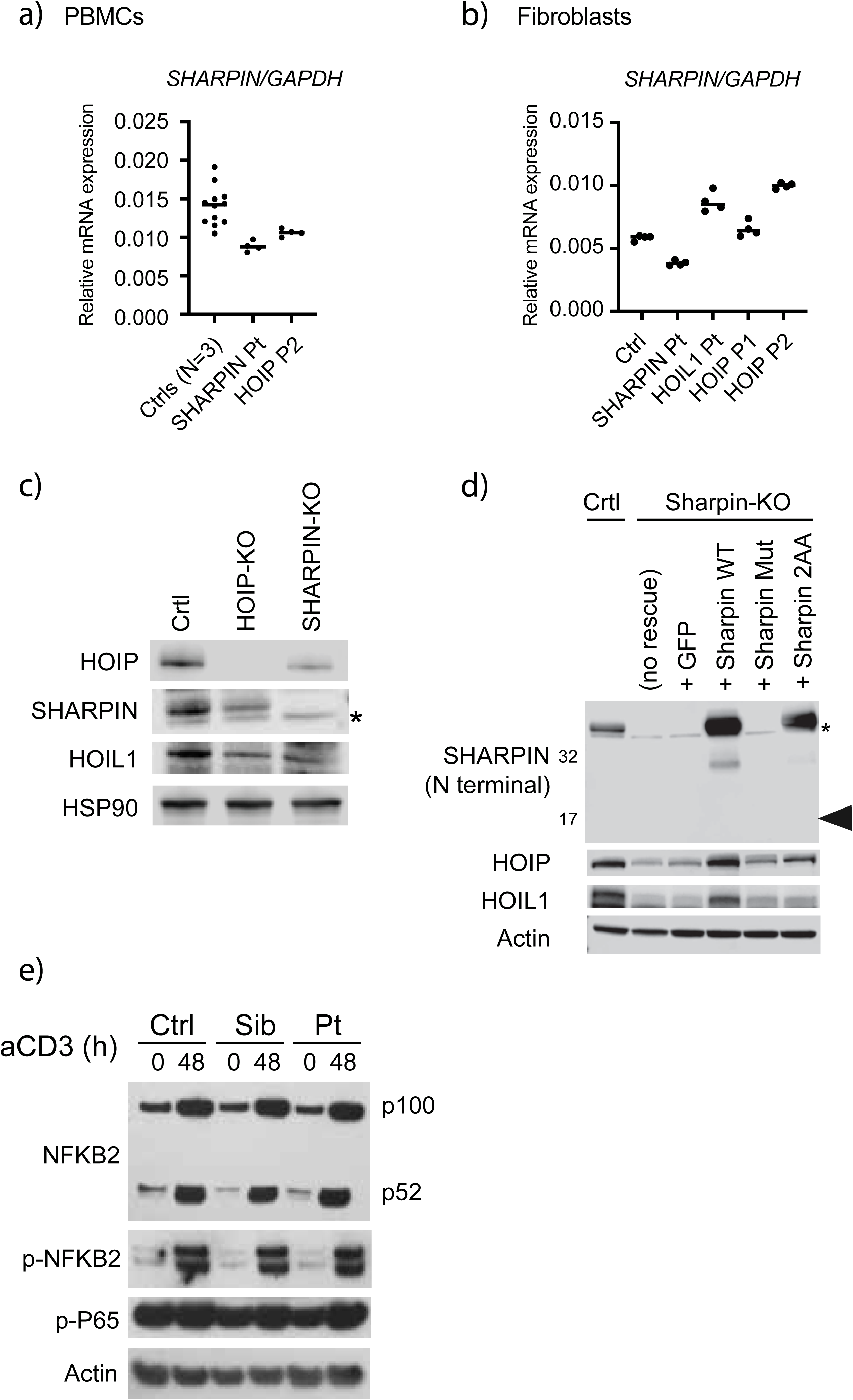
LUBAC destabilization and non-canonical NF-κ B activation in SHARPIN deficiency *in vitro* and *ex vivo.* (a) Normalized mRNA levels of SHARPIN in (a) PBMCs and (b) fibroblasts from LUBAC-deficient individuals and healthy controls. (c) Western blot confirmation of CRISPR-Cas9 mediated knockout of SHARPIN and HOIP in HeLa clones. * indicates a non-specific band. (d) Retroviral reconstitution of SHARPIN variants or GFP in a SHARPIN-KO HeLa clone. Mut: the frameshift variant observed in the patient (p.Leu74Profs*86); 2AA: a LUBAC-destabilizing positive control SHARPIN mutant (p.L179A/I183A) previously reported from structure-guided analysis ^19^. Note that the expected truncated SHARPIN protein (18kDa: arrowhead) is not observed. * indicates a non-specific band. (e) Normal induction of non-canonical NF-κB in SHARPIN deficient patient. Total PBMCs were stimulated with anti-CD3 (aCD3) for the indicated durations, and the expression of NFKB2 p100 (full length) and p52 (active form) was detected by western blot. Ctrl: unrelated healthy control; Sib: the patient’s sibling carrying a heterozygous SHARPIN frameshift variant; Pt: SHARPIN deficient patient.

**Extended Data Figure 3:**
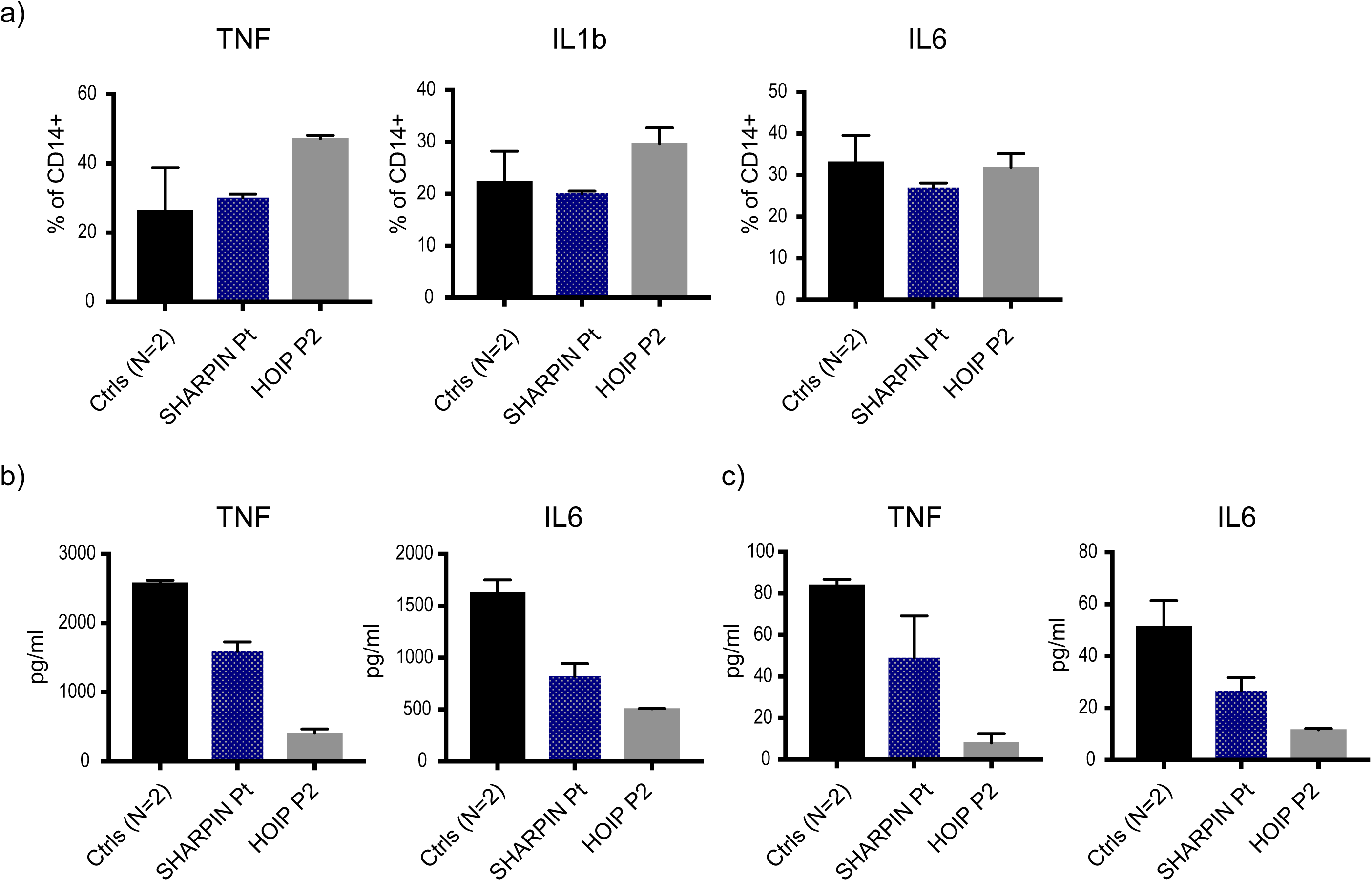
Cytokine expression studies *ex vivo* in LUBAC deficient patients. (a) Cytokine expression in LUBAC deficient monocytes. PBMCs from SHARPIN and HOIP deficient patients and two healthy controls were stimulated with IL1β (10 ng/ml) for 6h, and the intracellular accumulation of intracellular cytokines in CD14^+^ monocytes were quantified by flow cytometry. (b-c) Cytokine secretion from LUBAC deficient PBMCs. PBMCs from SHARPIN and HOIP deficient patients and two healthy controls were stimulated with (b) LPS (1 μg/ml) or (c) IL1β (10 ng/ml) for 6h, and secreted cytokines were measured by ELISA. All the experiments were performed with biological triplicates. A representative result of two independent experiments is shown.

**Extended Data Figure 4:**
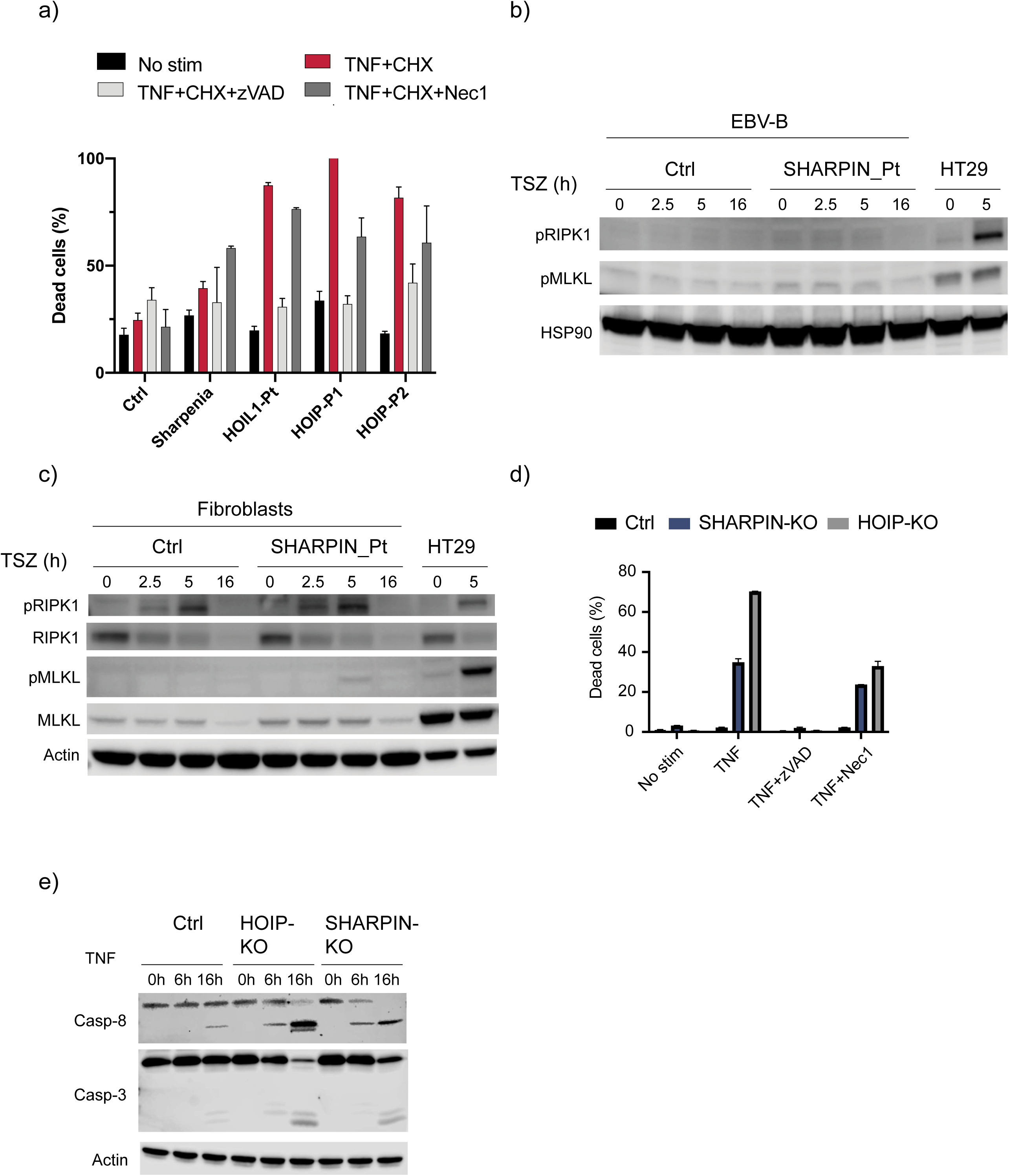
Cell death induction assays in human LUBAC deficiency. (a) Time-lapse quantification of dead cell percentages using fibroblasts from LUBAC deficient patients. The quantification was performed in biological triplicates. (b-c) Necroptosis induction assay using EBV-immortalized lymphoblastoid cells (b) and fibroblasts (c) from SHARPIN deficient patient. The fibroblasts were stimulated with TSZ (TNF, smac mimetic (compound A) and zVAD) for the indicated time. HT29 was used as a positive control for the phosphor-western blotting. (d) Time lapse quantification of dead cell percentages using SHARPIN- and HOIP- knockout HeLa cells. Dead cell quantification at 16 h time point was shown. The data acquisition was performed in biological triplicates. (e) Caspase cleavage assay using SHARPIN- and HOIP- knockout HeLa cells. (a-e) A representative result of more than three independent experiments is shown.

**Extended Data Figure 5:**
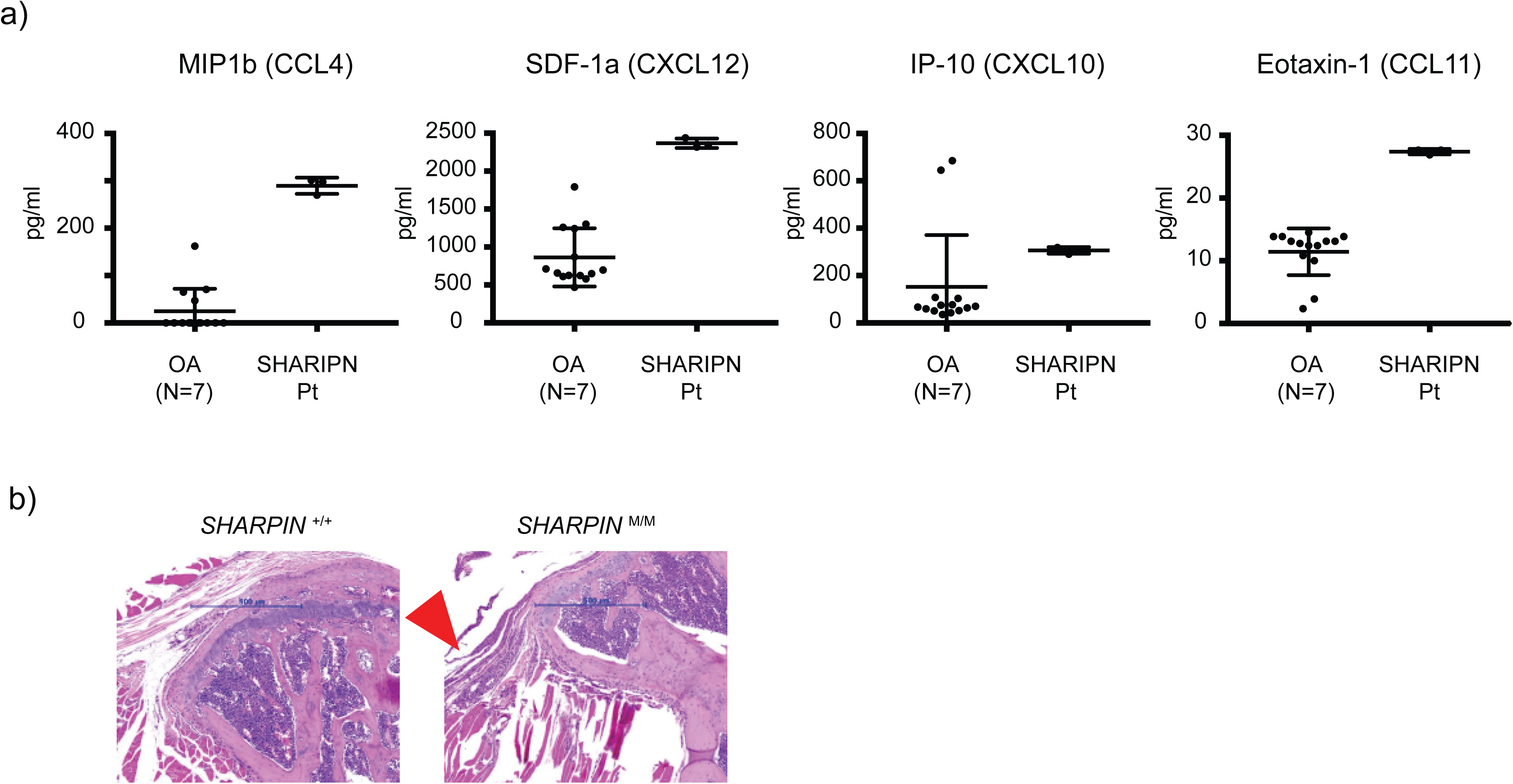
Characterization of joint inflammation in SHARPIN deficiency (a) Multiplex ELISA measurement of chemokines in the sterile synovial fluid of SHARPIN deficient patient before the initiation of anti-TNF treatment and osteoarthritis (OA) controls (N=7). The samples were measured in technical triplicate (the SHARPIN deficient patient) or in duplicate (OA controls), respectively. (b) Representative hematoxylin and eosin staining sections of elbow joints from *Sharpin* deficient mice and wild type littermate control. Arrowhead indicates inflamed ligament.

**Extended Data Figure 6:**
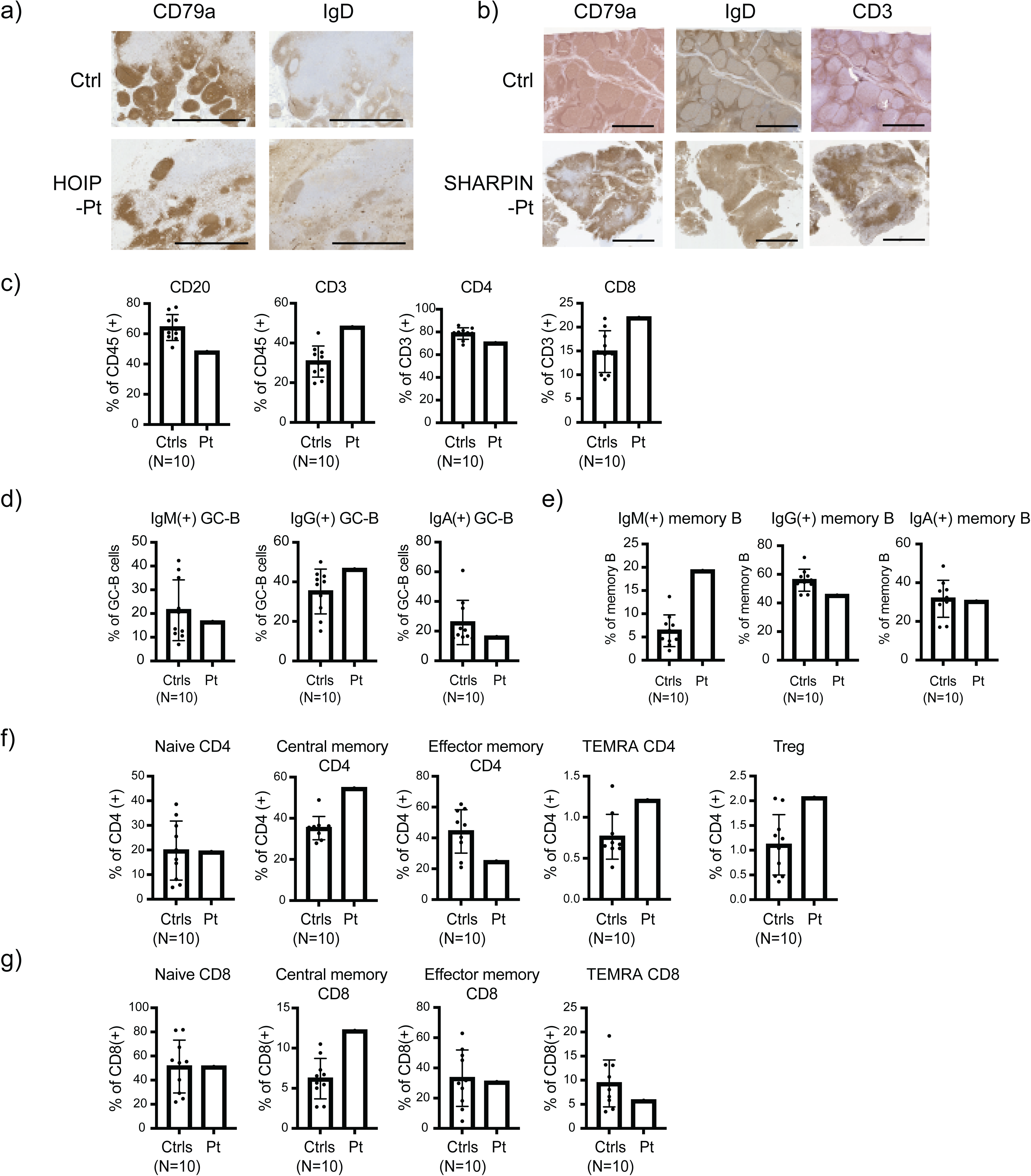
Characterization of secondary lymphoid organ abnormalities in LUBAC deficiencies. (a-b) Aberrant formation of lymphoid follicles and paracortex in secondary lymphoid organs from LUBAC deficient patients. (a) Lymph node histology of a HOIP deficient patient. (b) Adenoid histology of SHARPIN deficient patient. (c-g) High dimensional spectral flow cytometry analysis of human adenoid samples. (c) Quantification of CD3^+^, CD4^+^, CD8^+^ and CD20^+^ populations in adenoid samples. (d-e) Surface immunoglobulin expression in GC-B (d) and memory B (e) populations. (f-g) Quantification of T cell subpopulations in the adenoid samples.

**Extended Data Figure 7:**
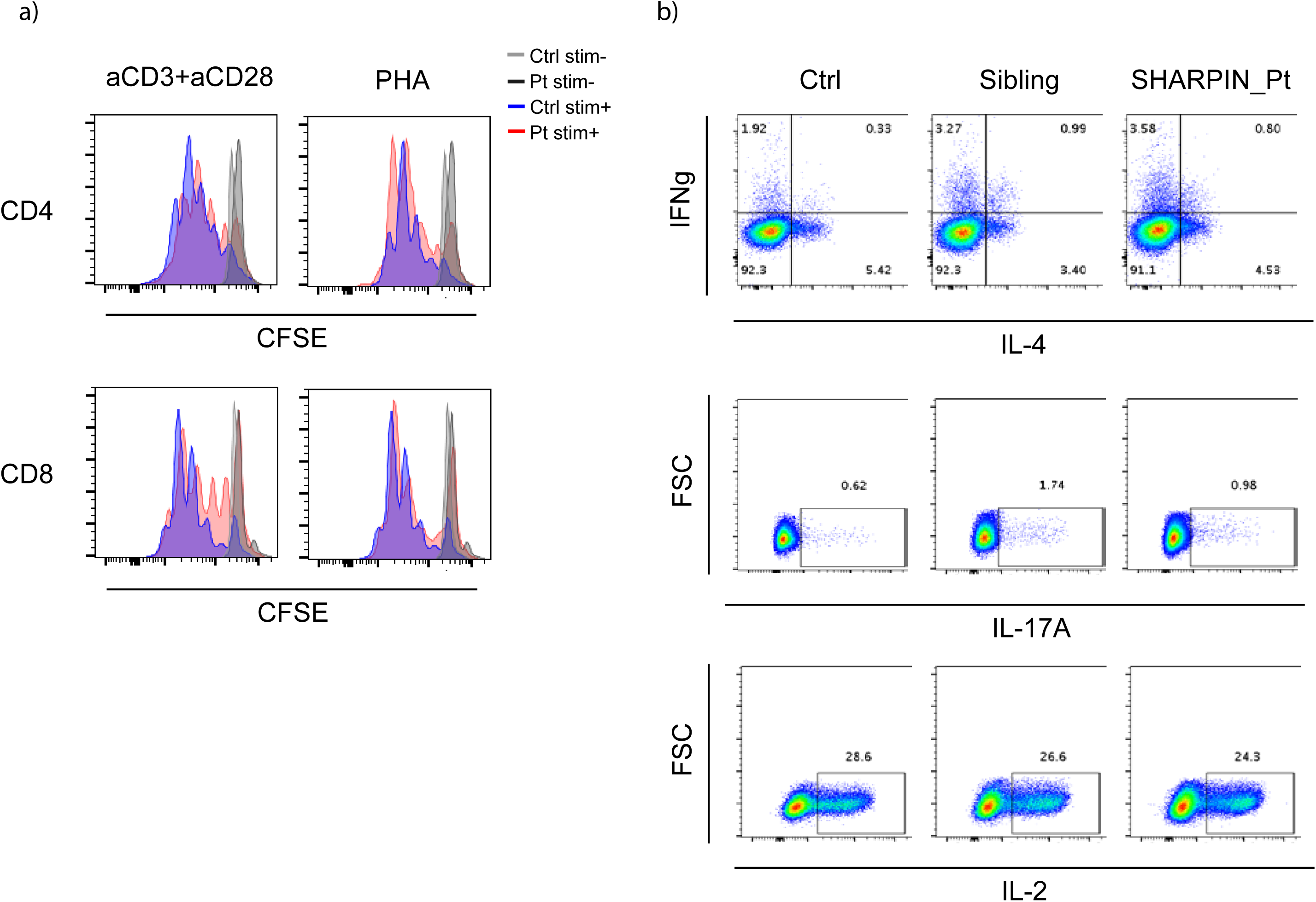
Normal T cell phenotyping results in the SHARPIN deficient patient *ex vivo*. (a) T cell proliferation assay. PMBCs were incubated with Cell Trave Violet, stimulated with anti-CD3/28 or PHA for 72h and analyzed by flow cytometer. (b) Intracellular cytokine staining for Th1, Th2 and Th17 populations. PBMCs were stimulated with PMA (100 ng/ml) and ionomycin (1 μM) for 5h with Brefeldin A. Stimulated cells were surface stained, fixed and permeabilized with BD Cytofix/Cytoperm kit. Cells were further stained for intracellular cytokines and analyzed by flow cytometry. Ctrl: unrelated healthy control, Sib: sibling carrying the heterozygous frameshift SHARPIN variant.

**Extended Data Figure 8:**
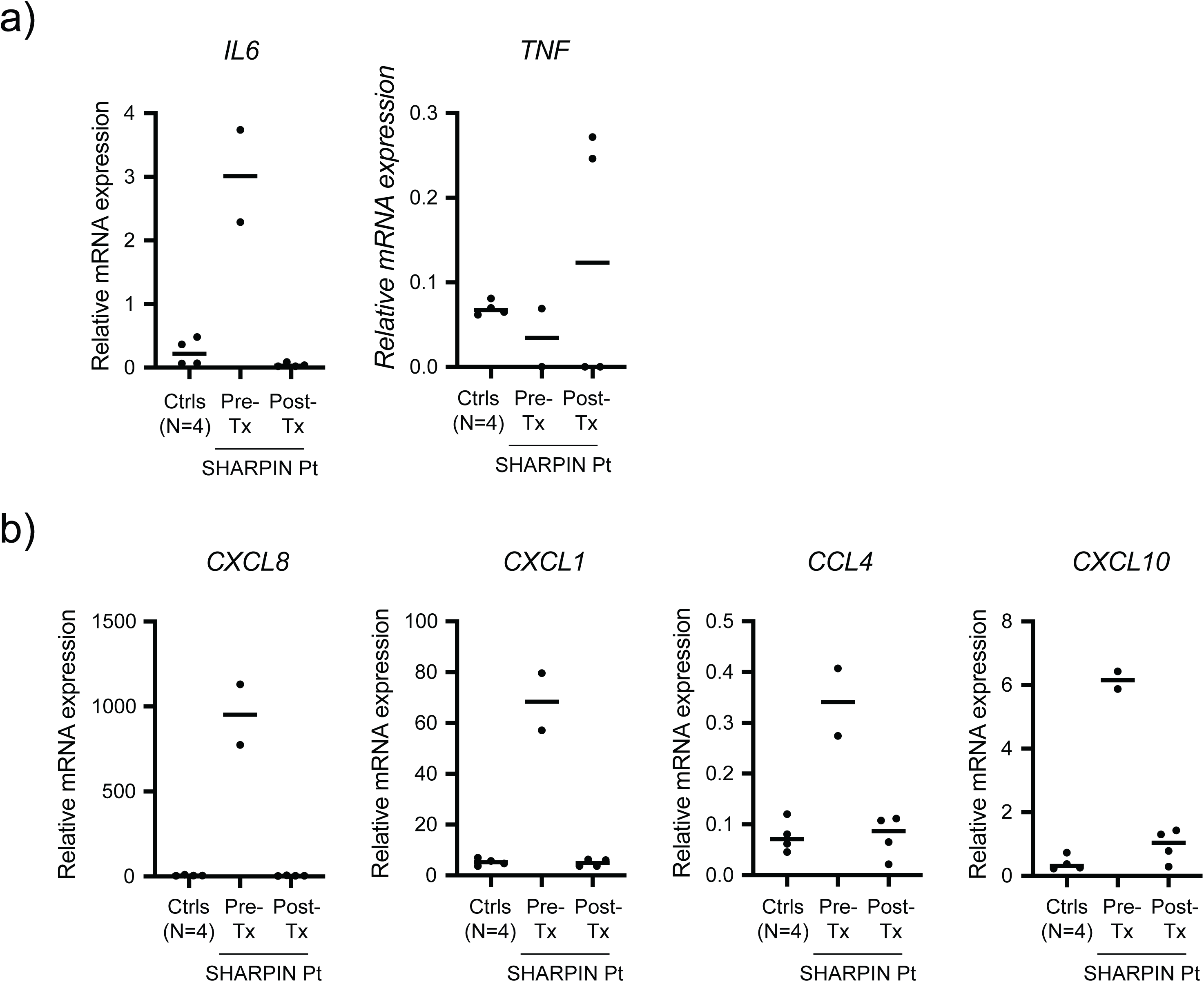
Whole blood RNA sequencing. (a-b) mRNA expression of selected cytokines (a) and chemokines (b) in the pre- and post-treatment SHARPIN deficient patient whole blood RNA samples as well as four age-matched healthy controls.

## REFERENCES

1. Oda, H. & Kastner, D. L. Genomics, Biology, and Human Illness: Advances in the Monogenic Autoinflammatory Diseases. Rheum Dis Clin North Am 43, 327–345 (2017). https://doi.org:10.1016/j.rdc.2017.04.011

2. Manthiram, K., Zhou, Q., Aksentijevich, I. & Kastner, D. L. The monogenic autoinflammatory diseases define new pathways in human innate immunity and inflammation. Nat Immunol 18, 832–842 (2017). https://doi.org:10.1038/ni.3777

3. Anderton, H., Wicks, I. P. & Silke, J. Cell death in chronic inflammation: breaking the cycle to treat rheumatic disease. Nat Rev Rheumatol 16, 496–513 (2020). https://doi.org:10.1038/s41584-020-0455-8

4. Iwai, K., Fujita, H. & Sasaki, Y. Linear ubiquitin chains: NF-kappaB signalling, cell death and beyond. Nat Rev Mol Cell Biol 15, 503–508 (2014). https://doi.org:10.1038/nrm3836

5. Spit, M., Rieser, E. & Walczak, H. Linear ubiquitination at a glance. J Cell Sci 132 (2019). https://doi.org:10.1242/jcs.208512

6. Fiil, B. K. & Gyrd-Hansen, M. The Met1-linked ubiquitin machinery in inflammation and infection. Cell Death Differ 28, 557–569 (2021). https://doi.org:10.1038/s41418-020-00702-x

7. Fuseya, Y. et al. The HOIL-1L ligase modulates immune signalling and cell death via monoubiquitination of LUBAC. Nat Cell Biol 22, 663–673 (2020). https://doi.org:10.1038/s41556-020-0517-9

8. Kelsall, I. R., Zhang, J., Knebel, A., Arthur, J. S. C. & Cohen, P. The E3 ligase HOIL-1 catalyses ester bond formation between ubiquitin and components of the Myddosome in mammalian cells. Proc Natl Acad Sci U S A 116, 13293–13298 (2019). https://doi.org:10.1073/pnas.1905873116

9. Boisson, B. et al. Immunodeficiency, autoinflammation and amylopectinosis in humans with inherited HOIL-1 and LUBAC deficiency. Nat Immunol 13, 1178–1186 (2012). https://doi.org:10.1038/ni.2457

10. Boisson, B. et al. Human HOIP and LUBAC deficiency underlies autoinflammation, immunodeficiency, amylopectinosis, and lymphangiectasia. J Exp Med 212, 939–951 (2015). https://doi.org:10.1084/jem.20141130

11. Oda, H. et al. Second Case of HOIP Deficiency Expands Clinical Features and Defines Inflammatory Transcriptome Regulated by LUBAC. Front Immunol 10, 479 (2019). https://doi.org:10.3389/fimmu.2019.00479

12. Gerlach, B. et al. Linear ubiquitination prevents inflammation and regulates immune signalling. Nature 471, 591–596 (2011). https://doi.org:10.1038/nature09816

13. Tokunaga, F. et al. SHARPIN is a component of the NF-kappaB-activating linear ubiquitin chain assembly complex. Nature 471, 633–636 (2011). https://doi.org:10.1038/nature09815

14. Ikeda, F. et al. SHARPIN forms a linear ubiquitin ligase complex regulating NF-kappaB activity and apoptosis. Nature 471, 637–641 (2011). https://doi.org:10.1038/nature09814

15. Peltzer, N. et al. LUBAC is essential for embryogenesis by preventing cell death and enabling haematopoiesis. Nature 557, 112–117 (2018). https://doi.org:10.1038/s41586-018-0064-8

16. Kumari, S. et al. Sharpin prevents skin inflammation by inhibiting TNFR1-induced keratinocyte apoptosis. Elife 3 (2014). https://doi.org:10.7554/eLife.03422

17. Rickard, J. A. et al. TNFR1-dependent cell death drives inflammation in Sharpin-deficient mice. Elife 3 (2014). https://doi.org:10.7554/eLife.03464

18. Peltzer, N. et al. HOIP deficiency causes embryonic lethality by aberrant TNFR1- mediated endothelial cell death. Cell Rep 9, 153–165 (2014). https://doi.org:10.1016/j.celrep.2014.08.066

19. Fujita, H. et al. Cooperative Domain Formation by Homologous Motifs in HOIL-1L and SHARPIN Plays A Crucial Role in LUBAC Stabilization. Cell Rep 23, 1192–1204 (2018). https://doi.org:10.1016/j.celrep.2018.03.112

20. Chien, S. J., Silva, K. A., Kennedy, V. E., HogenEsch, H. & Sundberg, J. P. The pathogenesis of chronic eosinophilic esophagitis in SHARPIN-deficient mice. Exp Mol Pathol 99, 460–467 (2015). https://doi.org:10.1016/j.yexmp.2015.08.012

21. Park, Y. et al. SHARPIN controls regulatory T cells by negatively modulating the T cell antigen receptor complex. Nat Immunol 17, 286–296 (2016). https://doi.org:10.1038/ni.3352

22. Casanova, J. L., Conley, M. E., Seligman, S. J., Abel, L. & Notarangelo, L. D. Guidelines for genetic studies in single patients: lessons from primary immunodeficiencies. J Exp Med 211, 2137–2149 (2014). https://doi.org:10.1084/jem.20140520

23. Karczewski, K. J. et al. The mutational constraint spectrum quantified from variation in 141,456 humans. Nature 581, 434–443 (2020). https://doi.org:10.1038/s41586-020-2308-7

24. Seymour, R. E. et al. Spontaneous mutations in the mouse Sharpin gene result in multiorgan inflammation, immune system dysregulation and dermatitis. Genes Immun 8, 416–421 (2007). https://doi.org:10.1038/sj.gene.6364403

25. Wijbrandts, C. A. et al. The clinical response to infliximab in rheumatoid arthritis is in part dependent on pretreatment tumour necrosis factor alpha expression in the synovium. Ann Rheum Dis 67, 1139–1144 (2008). https://doi.org:10.1136/ard.2007.080440

26. HogenEsch, H. et al. A spontaneous mutation characterized by chronic proliferative dermatitis in C57BL mice. Am J Pathol 143, 972–982 (1993).

27. Sundberg, J. P. et al. Keratinocyte-specific deletion of SHARPIN induces atopic dermatitis-like inflammation in mice. PLoS One 15, e0235295 (2020). https://doi.org:10.1371/journal.pone.0235295

28. Wang, J. et al. LUBAC Suppresses IL-21-Induced Apoptosis in CD40-Activated Murine B Cells and Promotes Germinal Center B Cell Survival and the T-Dependent Antibody Response. Front Immunol 12, 658048 (2021). https://doi.org:10.3389/fimmu.2021.658048

29. Roco, J. A. et al. Class-Switch Recombination Occurs Infrequently in Germinal Centers. Immunity 51, 337–350 e337 (2019). https://doi.org:10.1016/j.immuni.2019.07.001

30. Elsner, R. A. & Shlomchik, M. J. Germinal Center and Extrafollicular B Cell Responses in Vaccination, Immunity, and Autoimmunity. Immunity 53, 1136–1150 (2020). https://doi.org:10.1016/j.immuni.2020.11.006

31. Teh, C. E. et al. Linear ubiquitin chain assembly complex coordinates late thymic T-cell differentiation and regulatory T-cell homeostasis. Nat Commun 7, 13353 (2016). https://doi.org:10.1038/ncomms13353

32. HogenEsch, H. et al. Increased expression of type 2 cytokines in chronic proliferative dermatitis (cpdm) mutant mice and resolution of inflammation following treatment with IL-12. Eur J Immunol 31, 734–742 (2001). https://doi.org:10.1002/1521-4141(200103)31:3<734::aid-immu734>3.0.co;2-9

33. van Zelm, M. C. et al. Human CD19 and CD40L deficiencies impair antibody selection and differentially affect somatic hypermutation. J Allergy Clin Immunol 134, 135–144 (2014). https://doi.org:10.1016/j.jaci.2013.11.015

34. Meyers, G. et al. Activation-induced cytidine deaminase (AID) is required for B-cell tolerance in humans. Proc Natl Acad Sci U S A 108, 11554–11559 (2011). https://doi.org:10.1073/pnas.1102600108

35. McGowan, H. W. et al. Sharpin is a key regulator of skeletal homeostasis in a TNF-dependent manner. J Musculoskelet Neuronal Interact 14, 454–463 (2014).

36. Douglas, T., Champagne, C., Morizot, A., Lapointe, J. M. & Saleh, M. The Inflammatory Caspases-1 and -11 Mediate the Pathogenesis of Dermatitis in Sharpin-Deficient Mice. J Immunol 195, 2365–2373 (2015). https://doi.org:10.4049/jimmunol.1500542

37. Gurung, P., Sharma, B. R. & Kanneganti, T. D. Distinct role of IL-1beta in instigating disease in Sharpin(cpdm) mice. Sci Rep 6, 36634 (2016). https://doi.org:10.1038/srep36634

38. Gijbels, M. J., HogenEsch, H., Bruijnzeel, P. L., Elliott, G. R. & Zurcher, C. Maintenance of donor phenotype after full-thickness skin transplantation from mice with chronic proliferative dermatitis (cpdm/cpdm) to C57BL/Ka and nude mice and vice versa. J Invest Dermatol 105, 769–773 (1995). https://doi.org:10.1111/1523-1747.ep12325599

39. Anderton, H. et al. Langerhans cells are an essential cellular intermediary in chronic dermatitis. Cell Rep 39, 110922 (2022). https://doi.org:10.1016/j.celrep.2022.110922

40. Pasparakis, M. & Vandenabeele, P. Necroptosis and its role in inflammation. Nature 517, 311–320 (2015). https://doi.org:10.1038/nature14191

41. Mifflin, L., Ofengeim, D. & Yuan, J. Receptor-interacting protein kinase 1 (RIPK1) as a therapeutic target. Nat Rev Drug Discov 19, 553–571 (2020). https://doi.org:10.1038/s41573-020-0071-y

42. Badran, Y. R. et al. Human RELA haploinsufficiency results in autosomal-dominant chronic mucocutaneous ulceration. J Exp Med 214, 1937–1947 (2017). https://doi.org:10.1084/jem.20160724

43. Adeeb, F. et al. A Novel RELA Truncating Mutation in a Familial Behcet’s Disease-like Mucocutaneous Ulcerative Condition. Arthritis Rheumatol 73, 490–497 (2021). https://doi.org:10.1002/art.41531

44. Damgaard, R. B. et al. OTULIN deficiency in ORAS causes cell type-specific LUBAC degradation, dysregulated TNF signalling and cell death. EMBO Mol Med 11 (2019). https://doi.org:10.15252/emmm.201809324

45. Li, Y. et al. Human RIPK1 deficiency causes combined immunodeficiency and inflammatory bowel diseases. Proc Natl Acad Sci U S A 116, 970–975 (2019). https://doi.org:10.1073/pnas.1813582116

46. Cuchet-Lourenco, D. et al. Biallelic RIPK1 mutations in humans cause severe immunodeficiency, arthritis, and intestinal inflammation. Science 361, 810–813 (2018). https://doi.org:10.1126/science.aar2641

47. Lalaoui, N. et al. Mutations that prevent caspase cleavage of RIPK1 cause autoinflammatory disease. Nature 577, 103–108 (2020). https://doi.org:10.1038/s41586-019-1828-5

48. Tao, P. et al. A dominant autoinflammatory disease caused by non-cleavable variants of RIPK1. Nature 577, 109–114 (2020). https://doi.org:10.1038/s41586-019-1830-y

49. Taft, J. et al. Human TBK1 deficiency leads to autoinflammation driven by TNF-induced cell death. Cell 184, 4447–4463 e4420 (2021). https://doi.org:10.1016/j.cell.2021.07.026

50. Zhou, Q. et al. Biallelic hypomorphic mutations in a linear deubiquitinase define otulipenia, an early-onset autoinflammatory disease. Proc Natl Acad Sci U S A 113, 10127–10132 (2016). https://doi.org:10.1073/pnas.1612594113

51. Zinngrebe, J. et al. --LUBAC deficiency perturbs TLR3 signaling to cause immunodeficiency and autoinflammation. J Exp Med 213, 2671–2689 (2016). https://doi.org:10.1084/jem.20160041

52. Zhang, Q. et al. Inborn errors of type I IFN immunity in patients with life-threatening COVID-19. Science 370 (2020). https://doi.org:10.1126/science.abd4570

53. Schmitt, N. et al. IL-12 receptor beta1 deficiency alters in vivo T follicular helper cell response in humans. Blood 121, 3375–3385 (2013). https://doi.org:10.1182/blood-2012-08-448902

54. Kelsall, I. R. et al. HOIL-1 ubiquitin ligase activity targets unbranched glucosaccharides and is required to prevent polyglucosan accumulation. EMBO J 41, e109700 (2022). https://doi.org:10.15252/embj.2021109700

55. Otten, E. G. et al. Ubiquitylation of lipopolysaccharide by RNF213 during bacterial infection. Nature 594, 111–116 (2021). https://doi.org:10.1038/s41586-021-03566-4

56. Wang, K., Li, M. & Hakonarson, H. ANNOVAR: functional annotation of genetic variants from high-throughput sequencing data. Nucleic Acids Res 38, e164 (2010). https://doi.org:10.1093/nar/gkq603

57. Kim, D., Paggi, J. M., Park, C., Bennett, C. & Salzberg, S. L. Graph-based genome alignment and genotyping with HISAT2 and HISAT-genotype. Nat Biotechnol 37, 907–915 (2019). https://doi.org:10.1038/s41587-019-0201-4

58. Anders, S., Pyl, P. T. & Huber, W. HTSeq--a Python framework to work with high-throughput sequencing data. Bioinformatics 31, 166–169 (2015). https://doi.org:10.1093/bioinformatics/btu638

59. Robinson, M. D., McCarthy, D. J. & Smyth, G. K. edgeR: a Bioconductor package for differential expression analysis of digital gene expression data. Bioinformatics 26, 139–140 (2010). https://doi.org:10.1093/bioinformatics/btp616

60. Schneider, C. A., Rasband, W. S. & Eliceiri, K. W. NIH Image to ImageJ: 25 years of image analysis. Nat Methods 9, 671–675 (2012). https://doi.org:10.1038/nmeth.2089

61. Turchaninova, M. A. et al. High-quality full-length immunoglobulin profiling with unique molecular barcoding. Nat Protoc 11, 1599–1616 (2016). https://doi.org:10.1038/nprot.2016.093

62. Bolotin, D. A. et al. MiXCR: software for comprehensive adaptive immunity profiling. Nat Methods 12, 380–381 (2015). https://doi.org:10.1038/nmeth.3364

63. Shugay, M. et al. VDJtools: Unifying Post-analysis of T Cell Receptor Repertoires. PLoS Comput Biol 11, e1004503 (2015). https://doi.org:10.1371/journal.pcbi.1004503

