## Supplementary table 1-3 for "Human LUBAC deficiency leads to autoinflammation and immunodeficiency by dysregulation in TNF-mediated cell death"

### Supplementary Data Table 1. Peripheral blood lymphocyte surface markers

| | | SHARPIN Pt | | | | HOIP Pt | | | | Normal range | |
|---|---|---|---|---|---|---|---|---|---|---|---|
| | | **%** | | **Count** | | **%** | | **Count** | | **%** | **Count** |
| **Total T Cells** | CD3 | 80.4 | | 2307 | **High** | 77.9 | | 1799 | | 60.0-83.7 | 714-2266 |
| **T cell subsets** | CD4/CD3 | 51.1 | | 1467 | | 53.8 | | 1243 | | 31.9-62.2 | 359-1565 |
| | CD8/CD3 | 25.2 | | 723 | | 19 | | 439 | | 11.2-34.8 | 178-853 |
| | CD4/CD8 ratio | 2.03 | | | | 2.83 | | | | 1.11-5.17 | |
| | CD3+CD4-/CD8- | 3.1 | | 89 | | 4.8 | | 111 | | 1.3-9.2 | 18-185 |
| | CD4/CD3/CD62+/CD45RA+ | 15.2 | | 436 | | 28.8 | | 665 | | 7.6-37.7 | 102-1041 |
| | CD4/CD3/CD62+/CD45RA- | 27.4 | | 788 | **High** | 17 | | 393 | | 10.4-30.7 | 162-614 |
| | CD4/CD3/CD62-CD45RA- | 8.4 | | 241 | **High** | 5.4 | | 125 | | 2.3-15.6 | 42-225 |
| | CD4/CD3/CD62-CD45RA+ | 0.2 | | 6 | | 2.5 | **High** | 58 | **High** | 0-1.5 | 0-29 |
| | CD8/CD3/CD62+/CD45RA+ | 12.9 | | 370 | | 11.6 | | 268 | | 5.7-19.7 | 85-568 |
| | CD8/CD3/CD62+/CD45RA- | 6.2 | | 178 | | 3.1 | | 72 | | 1.5-10.3 | 25-180 |
| | CD8/CD3/CD62-/CD45RA- | 3.7 | | 106 | | 2.4 | | 55 | | 1.1-9.2 | 24-175 |
| | CD8/CD3/CD62-/CD45RA+ | 2.3 | | 66 | | 1.9 | | 44 | | 0.7-7.8 | 11-172 |
| | CD3/CD4/CD31/CD45RA | 8.7 | | 251 | | 25.7 | **High** | 593 | **High** | 1.6-20.2 | 34-426 |
| | CD3/CD4/CD45RA-/CXCR5 | 4.8 | | 138 | | 4.5 | | 104 | | 1.8-8.9 | 35-172 |
| | CD3/CD8/CD57 | NA | | NA | | 3.2 | | 74 | | <16.2 | <397 |
| | CD3/CD4/CD25/FoxP3 | 1.9 | | 55 | | 1 | **Low** | 23 | **Low** | 1.6-4.3 | 25-89 |
| **B cell subsets** | CD20 | 13.3 | | 382 | **High** | 17.5 | | 404 | **High** | 3.0-19.0 | 59-329 |
| | CD19 | 13.3 | | 382 | **High** | 17.5 | | 404 | **High** | 3.3-19.3 | 61-321 |
| | CD20/CD27 | 2.1 | | 60 | | 0.2 | **Low** | 5 | **Low** | 0.8-3.6 | 13-68 |
| | CD20/CD38 | 13.1 | | 377 | **High** | 16.9 | | 390 | **High** | 1.2-17.6 | 30-282 |
| | CD20/CD10 | 7.7 | **High** | 220 | **High** | 4.3 | | 100 | | 0.6-6.0 | 13-127 |
| | CD20/IgM-/CD38++ | 0.1 | | 3 | | 0 | | 0 | | 0-0.1 | 0-2 |
| | CD20/IgM+/CD10+ | 2.9 | **High** | 83 | **High** | 2.5 | | 58 | | 0.1-2.8 | 2.0-60 |
| | CD20/CD38+/CD10+ | 3 | **High** | 86 | **High** | 2.4 | | 55 | | 0.1-2.7 | 0-63 |
| | CD20/CD27+/IgM+ | 0.6 | | 17 | | 0.1 | **Low** | 2 | **Low** | 0.3-2.5 | 6.0-50 |
| | CD20/CD27+/IgM- | 1.3 | | 37 | | 0.1 | **Low** | 2 | **Low** | 0.3-2.2 | 6.0-43 |
| | CD19/CD24hi/CD38hi | 2.9 | **High** | 83 | **High** | 2.5 | **High** | 58 | **High** | 0.3-2.2 | 6.0-48 |
| | CD19/CD24-/CD38++ | 0.6 | **High** | 17 | **High** | 0.1 | | 2 | | 0-0.3 | 0-8 |
| | CD19/CD21low/CD38low | 0.1 | | 3 | | 0.1 | | 2 | | 0.1-0.8 | 1.0-16 |
| **NK Cells** | CD16+orCD56+/CD3- | 6 | **Low** | 172 | | 4.7 | **Low** | 109 | **Low** | 6.2-34.6 | 126-729 |
| | CD16+orCD56+/CD3+ | 10 | | 287 | | 6.5 | | 150 | | 2.2-12.4 | 29-299 |

#### Supplementary Data Table 2. Vaccine responses

| | Patient | | |
|---|---|---|---|
| **PCV-13*** | Pre | Post (4-weeks) | 2-fold increase |
| Serotype 1 | N/A** | N/A** | N/A** |
| Serotype 3 | 0.4 | 1.3 | (+) |
| Serotype 4 | 2.4 | 10.6 | (+) |
| Serotype 5 | 1.5 | 2.1 | (-) |
| Serotype 14 | 8.8 | 8.7 | (-) |
| Serotype 19F | 4.5 | 15.2 | (+) |
| Serotype 23F | 14.8 | 17.2 | (-) |
| Serotype 6B | 0.3 | 3.5 | (+) |
| Serotype 7F | 6.6 | 3.5 | (-) |
| Serotype 18C | 0.3 | 0.8 | (+) |
| Serotype 19A | 0.4 | 5.6 | (+) |
| Serotype 9V | 7.4 | 28.5 | (+) |
| **2-fold increase rate** | | | **63% (positive)***** |

* Note that serotype 6A was not included in the assessment.

** Serotype 1 was unable to quantitate due to technical reasons.

*** Antibody concentrations increased by 2-fold or greater for at least 50% of serotypes when comparing the pre- and post-vaccination results is considered positive.

| | Patient | Cutoff | |
|---|---|---|---|
| **VZV IgG (index)** | 343 | 165 | **Positive** |
| **HiB IgG (mg/L)** | >9.00 | 0.15 | **Positive** |
| **Diphtheria IgG (IU/ml)** | 0.03 | 0.01 | **Positive** |
| **Tetanus IgG (IU/ml)** | 0.48 | 0.01 | **Positive** |

Supplementary Data Table 3: inborn errors of cell death (IECD)

| | Autoinflammation | Immunodeficiency | Immunophenotype | Cell Death | Mouse phenotype | Treatment |
|---|---|---|---|---|---|---|
| Sharpenia | Fever<br>Arthritis<br>GI inflammation<br>Growth failure | Chronic otitis media by GAS | Abnormalities in germinal center development | Apoptosis<br>(> Necroptosis) | Dermatitis | Anti-TNF |
| HOIP deficiency | Fever<br>Arthritis<br>GI inflammation<br>Dermatitis<br>Growth failure | Viral encephalitis<br>Skin abscesses<br>Sepsis | Hypogammaglobulinemia<br>Low T cells<br>Low memory B cells<br>Vaccine failure | Apoptosis<br>(> Necroptosis) | Embryonic lethal | Anti-TNF<br>IVIG |
| HOIL1 deficiency | Fever<br>GI inflammation<br>Dermatitis<br>Growth failure | Sepsis<br>Meningitis | Hypogammaglobulinemia<br>Low memory B cells<br>Vaccine failure | Apoptosis<br>(> Necroptosis) | Embryonic lethal | Corticosteroids<br>HSCT |
| RELA haploinsufficiency | Fever<br>GI inflammation<br>Dermatitis<br>Arthritis | No | (*) | Apoptosis | Exacerbation in inducible models (**) | Anti-TNF |
| RIPK1 deficiency | Fever<br>Dermatitis<br>GI inflammation<br>Growth failure | Sepsis<br>Pneumonia<br>Skin abscesses<br>UTI | Low T cells<br>Low memory B cells | Apoptosis and necroptosis | Neonatal lethal | Anti-TNF<br>HSCT |
| CRIA | Fever<br>Lymphadenopathy<br>GI inflammation | No | No major abnormalities | Apoptosis and necroptosis | Embryonic lethal in homozygous knock-in mice | Anti-IL6 |

| | | | | | | |
|---|---|---|---|---|---|---|
| Otulipenia/ORAS | Fever<br>Panniculitis<br>GI inflammation<br>Sterile abscess formation<br>Growth failure | (***) | No major abnormalities | Apoptosis | Embryonic lethal | Anti-TNF<br>HSCT |
| TBK1 deficiency | Fever<br>Arthritis<br>Vasculitis<br>Basal ganglia calcification<br>Growth failure | No | No major abnormalities | Necroptosis (>apoptosis) | Embryonic lethal | Anti-TNF |

* One patient is noted to have $CD4^+$ T lymphoproliferation. ** TNF-induced dermatitis and dextran sulfate sodium (DSS)-induced colitis. *** Susceptibility to staphylococcal infections in haploinsufficient carriers of *OTULIN* mutations. Abbreviations: GAS: group A streptococcus; CRIA: cleavage-resistant RIPK1-induced autoinflammation; ORAS: otulin-related autoinflammatory syndrome; HSCT: hematopoietic stem cell transplantation, UTI: urinary tract infection.
